# Mapping disease loci to biological processes via joint pleiotropic and epigenomic partitioning

**DOI:** 10.1101/2025.05.05.25327017

**Authors:** Gaspard Kerner, Nolan Kamitaki, Benjamin Strober, Alkes L. Price

## Abstract

Genome-wide association studies (GWAS) have identified thousands of disease-associated loci, yet their interpretation remains limited by the heterogeneity of underlying biological processes. We propose Joint Pleiotropic and Epigenomic Partitioning (J-PEP), a clustering framework that integrates pleiotropic SNP effects on auxiliary traits and tissue-specific epigenomic data to partition disease-associated loci into biologically distinct clusters. To benchmark J-PEP against existing methods, we introduce a metric—Pleiotropic and Epigenomic Prediction Accuracy (PEPA)—that evaluates how well the clusters predict SNP-to-trait and SNP-to-tissue associations using off-chromosome data, avoiding overfitting. Applying J-PEP to GWAS summary statistics for 165 diseases/traits (average *N*=290K), we attained 16-30% higher PEPA than pleiotropic or epigenomic partitioning approaches with larger improvements for well-powered traits, consistent with simulations; these gains arise from J-PEP’s tendency to upweight correlated structure—signals present in both auxiliary trait and tissue data—thereby emphasizing shared components. For type 2 diabetes (T2D), J-PEP identified clusters refining canonical pathological processes while revealing underexplored immune and developmental signals. For hypertension (HTN), J-PEP identified stromal and adrenal-endocrine processes that were not identified in prior analyses. For neutrophil count, J-PEP identified hematopoietic, hepatic-inflammatory, and neuroimmune processes, expanding biological interpretation beyond classical immune regulation. Notably, integrating single-cell chromatin accessibility data refined bulk-based clusters, enhancing cell-type resolution and specificity. For T2D, single-cell data refined a bulk endocrine cluster to pancreatic islet β-cells, consistent with established β-cell dysfunction in insulin deficiency; for HTN, single-cell data refined a bulk endocrine cluster to adrenal cortex cells, consistent with a GO enrichment for neutrophil-mediated inflammation that implicates feedback between aldosterone production in the adrenal gland and local immune signaling. In conclusion, J-PEP provides a principled framework for partitioning GWAS loci into interpretable, tissue-informed clusters that provide biological insights on complex disease.

## Introduction

A central challenge in complex disease genomics is to decipher the biological processes underlying the thousands of loci identified by genome-wide association studies (GWAS)^1–6^. While interpretation of individual loci has yielded important insights into disease biology and functionally validated putative causal genes^7–11^, findings at individual loci remain limited^1,12^, motivating genome-wide approaches to map disease loci to biological processes. Extensive pleiotropy between diseases and complex traits^13–22^ motivates pleiotropic partitioning^23–25^ (also see refs. ^26–31^)—leveraging pleiotropic associations with auxiliary traits to group disease loci into clusters that often align with disease-critical cell types and biological pathways. However, interpreting clusters based solely on pleiotropy can be challenging when biological processes are partially overlapping or produce similar multi-trait signatures. Epigenomic annotations provide valuable information about disease biology^33–38^, and have been used to retrospectively interpret clustering results^23–25^, but have not been directly integrated into clustering models. Furthermore, no quantitative metric has been proposed to evaluate clustering performance.

Here, we introduce Joint Pleiotropic and Epigenomic Partitioning (J-PEP)—a method that jointly partitions disease loci into clusters using both pleiotropic associations with auxiliary traits and tissue-specific epigenomic profiles; J-PEP clusters capture both SNP-to-trait and SNP-to-tissue associations, enhancing cluster interpretability. We also introduce a new evaluation metric, Pleiotropic and Epigenomic Prediction Accuracy (PEPA), that evaluates how well the clusters predict SNP-to-trait and SNP-to-tissue associations. We compare J-PEP to pleiotropic and epigenomic partitioning approaches via simulations and analyses of 165 GWAS diseases/traits, using the PEPA metric. We find that J-PEP outperforms other approaches using the PEPA metric and we highlight novel biological insights for type 2 diabetes (T2D), hypertension (HTN) and neutrophil count (NC). Finally, we show that by integrating single-cell chromatin accessibility data, J-PEP enables the association of clusters—i.e., subsets of loci—with fine-grained cell types, refining broader bulk-derived associations.

## Results

### Overview of Methods

J-PEP partitions disease loci into biologically interpretable clusters by jointly leveraging pleiotropic data (across auxiliary traits) and epigenomic data (across tissues) (**Fig. 1**). For a given focal disease/trait, J-PEP inputs a SNP-to-trait matrix (*V*_*trait*_) derived from fine-mapping results^39,40^ across auxiliary traits, with each entry defined as the product of posterior inclusion probabilities (PIPs) for a given fine-mapped SNP (focal trait PIP >0.01) across the focal trait and a given auxiliary trait (**Methods**). To accommodate the sign of pleiotropic effects, each auxiliary trait is split into two components: one representing concordant (or positive) pleiotropic effects and the other representing discordant (or negative) pleiotropic effects (**Methods**). J-PEP also inputs a SNP-to-tissue matrix (*V*_*tissue*_) derived from epigenomic data across tissues, where each entry reflects the normalized strength of association between a fine-mapped SNP and a tissue, computed using an Expectation-Maximization (EM) algorithm (**Methods**). J-PEP then jointly factorizes *V*_*trait*_ and *V*_*tissue*_ to produce estimates of a SNP-to-cluster membership matrix (*W*), an auxiliary trait-to-cluster profile matrix (*H*_*trait*_), and a tissue-to-cluster profile matrix (*H*_*tissue*_), using an extended version of joint Bayesian non-negative matrix factorization (bNMF)^32^. Standard bNMF decomposes a single non-negative data matrix into lower-dimensional non-negative factors under Bayesian priors; J-PEP extends this framework to two matrices, encouraging cross-data alignment across clusters (see below). To enhance cluster interpretability, J-PEP restricts each tissue to a single cluster, whereas auxiliary traits may span multiple clusters. This iterative optimization process continues until convergence, yielding clusters that integrate pleiotropic and epigenomic information. To enhance stability, we perform ten optimization runs with different random seeds and select the run with the highest likelihood among those yielding the most consistent number of identified clusters (**Methods**). We note that of all input/output matrices, only *V*_*tissue*_ has normalized rows that specifically represent probabilities.

**Fig. 1.**
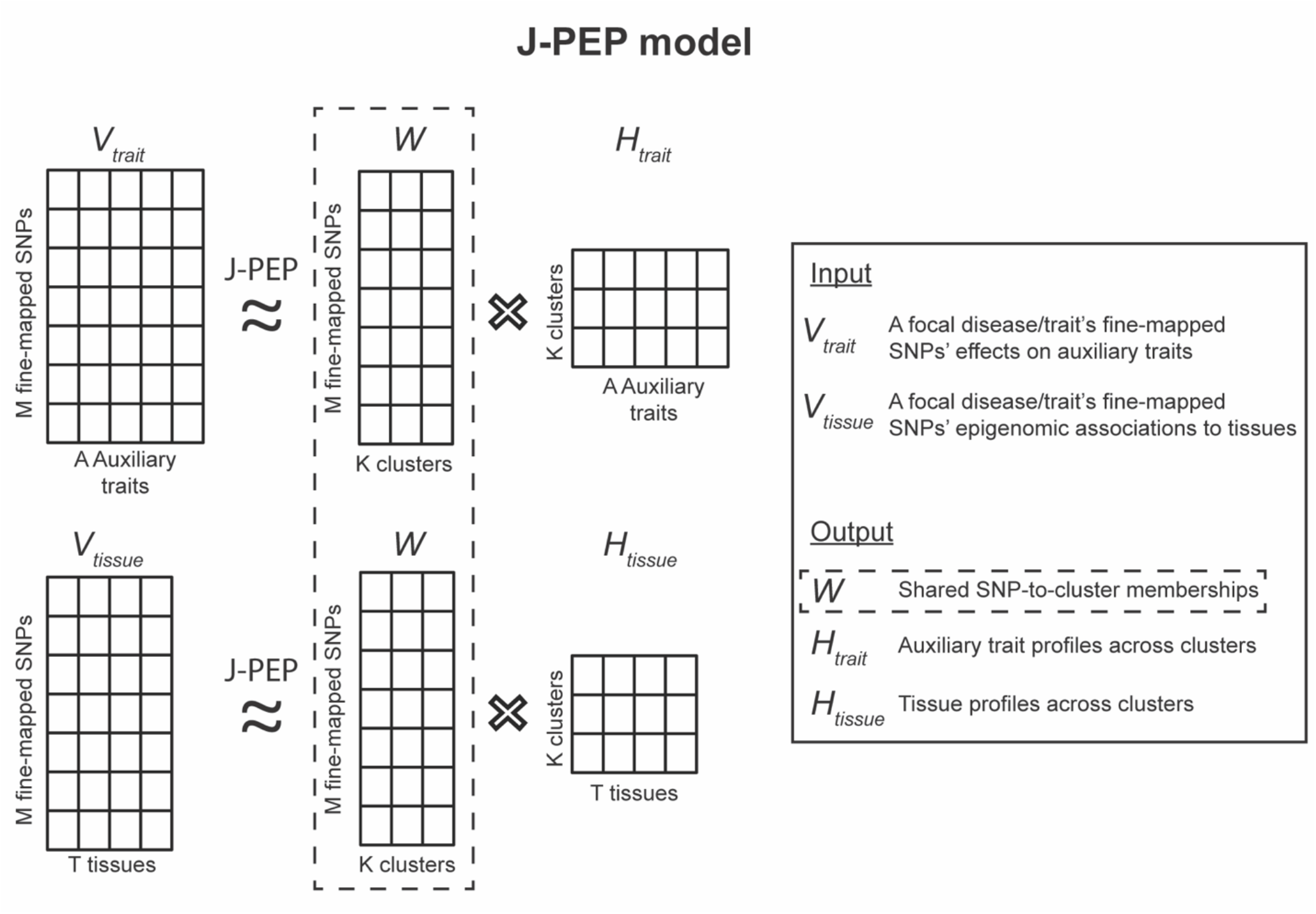
Schematic representation of the J-PEP model. The J-PEP model performs an extended version of joint Bayesian non-negative matrix factorization (bNMF) to fine-mapped SNPs from a given focal disease/trait. It jointly factorizes two input matrices: a SNP-to-trait matrix (*V*_*trait*_) and a SNP-to-tissue matrix (*V*_*tissue*_). J-PEP infers a shared SNP-to-cluster membership matrix (*W*), an auxiliary trait-to-cluster profile matrix (*H*_*trait*_) and a tissue-to-cluster profile matrix (*H*_*tissue*_).

In detail, J-PEP solves the following optimization problem:

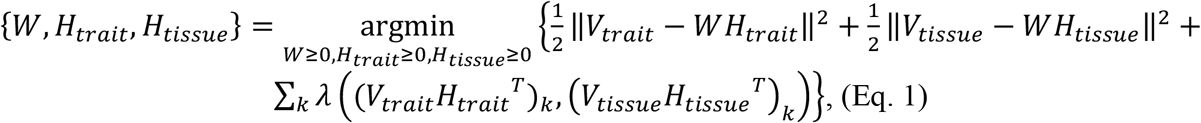

where *W, H*_*trait*_, *H*_*tissue*_, *V*_*trait*_ and *V*_*tissue*_ are as defined above, and (*V*_*trait*_*H*_*trait*_^*T*^)_*k*_ and (*V*_*tissue*_*H*_*tissue*_^*T*^)_*k*_ are vectors of length *M* (the number of focal disease/trait fine-mapped SNPs with PIP >0.01) representing the projections of fine-mapped SNPs onto the *k*^th^ cluster as defined by auxiliary traits and tissues, respectively. To guide the factorization process, the first two terms of the optimization function minimize the reconstruction error of the input matrices, while the third term is a penalization term that promotes alignment between pleiotropic and epigenomic data and suppresses irrelevant clusters. Specifically, *λ*(*A, B*) = −*log*_10_(*p*_*corr*_(*A, B*))^−1^, where *p*_*corr*_ indicates the one-sided Pearson correlation *p* value for positive correlation. Additionally, J-PEP enforces sparsity in tissue profiles by assigning each tissue to the cluster with the highest tissue value at each iteration of the algorithm. The initial number of clusters (set to 15 following prior work^23^) serves as an upper bound and may be reduced during optimization as uninformative clusters are pruned.

We compared J-PEP to two simpler approaches: Pleiotropic partitioning^23–25^, which factorizes a SNP-to-trait matrix (e.g. *V*_*trait*_) without incorporating tissue information; and Epigenomic partitioning, which factorizes a SNP-to-tissue matrix (e.g. *V*_*tissue*_) without incorporating pleiotropic information (**Methods**). In both approaches, partitioning is performed using standard bNMF. For Pleiotropic partitioning, we learn *H*_*tissue*_ from *V*_*tissue*_ and *W* using gradient descent by solving *V*_*tissue*_ = *W* × *H*_*tissue*_. For Epigenomic partitioning, we learn *H*_*trait*_ from *V*_*trait*_ And *W* using gradient descent by solving *V*_*trait*_ = *W* × *H*_*trait*_. To support comparisons between methods, the sparsity constraint on tissue profiles applied to J-PEP was also imposed on the two simpler approaches (**Methods**).

We applied J-PEP, Pleiotropic partitioning, and Epigenomic partitioning to GWAS summary statistics for 165 focal diseases/traits (average of 290K samples and 77 fine-mapped loci; a fine-mapped locus is defined as a locus containing at least one SNP with posterior inclusion probability (PIP) > 0.5), including 57 UK Biobank diseases/traits^41^ and 108 non-UK Biobank diseases/traits (**Supplementary Table 1**; see Data Availability). We integrated EpiMap data^36^ comprising 1,595 chromatin tracks, each defined by a unique combination of one of six histone or accessibility marks (DNase HS, H3K4me1, H3K4me2, H3K4me3, H3K9ac, H3K27ac) and a cell type label. These tracks are grouped into 32 broad tissue categories defined by EpiMap^36^ (**Methods**; see Data Availability); we refined our interpretation of J-PEP clusters using scATAC-seq^42^, profiling 615,998 nuclei across 30 adult human tissues, ultimately resolving 111 fine-grained cell types. We restricted most analyses to 55 approximately independent focal diseases/traits (pairwise *r*_*g*_ < 0.5), prioritizing traits with greater statistical power (defined by the number of fine-mapped loci); focal traits yielding fewer than two clusters for any of the three partitioning methods were excluded from most comparisons, yielding 38 focal diseases/traits (**Methods**). Auxiliary traits for each focal trait were selected from the full set of 165 diseases/traits based on two criteria: (1) a genetic correlation of < 0.5 with the focal trait, and (2) the existence of at least one shared causal SNP (PIP > 0.5) with the focal trait; on average, a focal disease/trait had 58 auxiliary traits. To improve cluster interpretability in selected real trait examples, we also conducted gene ontology enrichment^43^ informed by SNP-to-gene linking scores from ref. ^44^ (**Methods**).

To assess the performance of J-PEP and compare it to other clustering approaches, we developed Pleiotropic and Epigenomic Prediction Accuracy (PEPA), a new validation metric that evaluates how well a method predicts pleiotropic information (SNP-to-trait associations; *V*_*trait*_) from epigenomic information (SNP-to-tissue associations; *V*_*tissue*_) and vice versa. PEPA employs an off-chromosome prediction framework, where entries in *V*_*trait*_ (resp. *V*_*tissue*_) are predicted from the corresponding entries in *V*_*tissue*_ (resp. *V*_*trait*_) using cluster-specific profiles (*H*_*trait*_ And *H*_*tissue*_) estimated using off-chromosome data. We note that the off-chromosome prediction framework ensures that this metric is robust to overfitting. PEPA is defined as the geometric mean of two constituent metrics: Pleiotropic Prediction Accuracy (PPA), which measures how well SNP-to-trait associations (*V*_*trait*_) can be predicted from SNP-to-tissue associations (*V*_*tissue*_); and Epigenomic Prediction Accuracy (EPA), which measures how well SNP-to-tissue associations (*V*_*tissue*_) can be predicted from SNP-to-trait associations (*V*_*trait*_). Standard errors on each metric are computed using a genomic block-jackknife, similar to ref. ^35^. Further details are provided in the **Methods** section. We have released open-source software implementing J-PEP and PEPA (see Code Availability). We have released input and output of J-PEP, Pleiotropic partitioning and Epigenomic partitioning and PEPA results for the 165 diseases/traits (see Data Availability).

### Simulations

To evaluate the performance of different clustering methods, we performed simulations by simulating GWAS summary statistics at real SNPs, incorporating genetic architectures informed by predefined cluster profiles (**Methods**). Our primary simulations included 32 tissues and 20 auxiliary traits, all of which were genetically uncorrelated with one another. Briefly, we performed eight steps: (1) generate *H*_*trait*_ and *H*_*tissue*_ matrices representing cluster profiles (*K*_*causal*_ = 5 causal clusters with each cluster defined by a single auxiliary trait and a single tissue, in primary simulations); (2) construct the SNP-to-cluster membership matrix *W*, based on *H*_*tissue*_ and a SNP-to-tissue score matrix (similar to *V*_*tissue*_) obtained from real EpiMap data^36^ (see *Overview of Methods*) on all available SNPs from chromosomes 1 and 2 of the 1000 Genomes Project^45^; (3) define the SNP-to-trait association matrix *V*_*trait*_ = *WH*_*trait*_; (4) sample the identities of *m* = 100-400 causal SNPs (to evaluate different numbers of fine-mapped loci) from unlinked loci with probabilities proportional to the rows of *V*_*trait*_, i.e. their aggregated memberships across clusters (noting that the rows of *V*_*trait*_ are not normalized and thus do not represent probabilities); (5) sample standardized causal SNP effect sizes *β* for the focal disease/trait from a standard normal distribution, rescaling to achieve a total SNP-heritability of *h*^2^ = 025; (6) sample shared causal SNPs for the 20 auxiliary traits with probabilities proportional to the corresponding columns of *V*_*trait*_, enforcing a minimum of 25% overlap with the focal disease/trait for auxiliary traits with non-zero contributions to causal clusters; (7) compute GWAS summary statistics 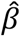using reference linkage disequilibrium (LD) matrices from the 1000 Genomes Project^45^ incorporating sampling noise consistent with the LD structure and the effective sample size of the reference panel (**Methods**); and (8) perform single causal variant fine-mapping^39,40^ for the focal disease/trait and auxiliary traits using the simulated GWAS summary statistics. As a result of steps (1) – (4), the SNP-to-trait and SNP-to-tissue association scores are not independent and are expected to exhibit non-zero correlations, reflecting shared structure imposed by the predefined cluster profiles. Our primary simulations also included uncorrelated structure, i.e. structure within *V*_*tissue*_ (resp. *V*_*trait*_) that is not correlated to any structure in *V*_*trait*_ (resp. *V*_*tissue*_) by injecting random subsets of *s* non-causal tissues and *s* non-causal traits with permuted profiles from causal ones; we also performed simulations with other parameter configurations (**Methods**). We note that uncorrelated structure is likely to arise in real data, e.g. if the relevant tissues or cell types driving pleiotropic associations to auxiliary traits are missing or underrepresented in epigenomic annotations, or analogously if a cell-type specific disease process is not captured by the available auxiliary traits.

To illustrate the interpretability of J-PEP relative to alternative methods, we first examined a representative simulation (**Fig. 2a**, Truth panel; we reduced the number of causal clusters in this simulation from 5 to 4 for easier visualization), with some auxiliary traits and tissues exhibiting uncorrelated structure (not shown; see **Methods**). Pleiotropic partitioning identified two clusters: a (purple) cluster spanning two causal tissues (one tissue with partial trait specificity, and one tissue with only spurious tissue-trait associations; top row), and a cluster spanning one causal tissue and two non-causal tissues (with only spurious tissue-trait associations in all three tissues). Epigenomic partitioning identified three clusters: a (green) cluster spanning one causal tissue with partial trait specificity, and two clusters each spanning one non-causal tissue (with only spurious trait-tissue associations). In contrast, J-PEP identified two clusters each spanning one causal tissue with partial trait specificity; J-PEP did not identify the remaining two clusters that were simulated. These results illustrate J-PEP’s ability to resolve trait–tissue relationships by leveraging shared structure across data modalities.

**Fig. 2.**
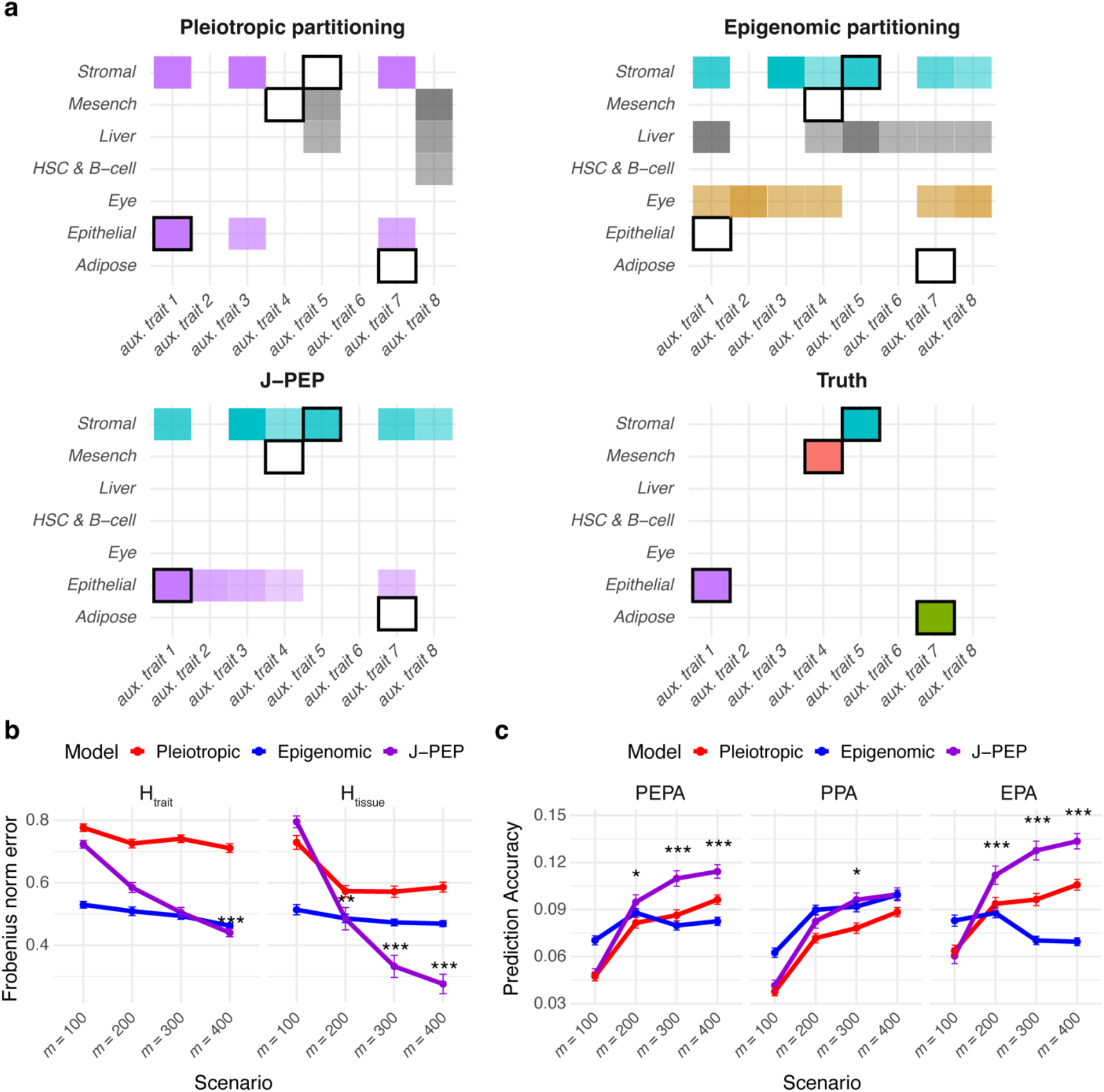
J-PEP outperforms Pleiotropic partitioning and Epigenomic partitioning in simulations. (a) Representative simulation illustrating the output of each method. Each color denotes one true or inferred cluster. Black squares denote causal tissue-auxiliary trait associations of true clusters. (b) Frobenius norm errors between the true vs. inferred auxiliary trait-to-cluster profile matrix (*H*_*trait*_) and tissue-to-cluster profile matrix (*H*_*tissue*_). (c) PEPA, PPA and EPA prediction accuracy metrics. In (b) and (c), results represent averages across 100 simulation replicates under four simulation settings with 100, 200, 300, or 400 causal SNPs for the focal disease/trait (denoted along the x-axis as *m* = 100, 200, 300, or 400, respectively). Error bars indicate standard errors across replicates. *: *p* < 0.05, **: *p* < 0.01, ***: *p* < 0.001. Numerical results are reported in **Supplementary Table 2**.

We evaluated the accuracy of Pleiotropic partitioning, Epigenomic partitioning and J-PEP in reconstructing the auxiliary trait-to-cluster profile matrix (*H*_*trait*_) and tissue-to-cluster profile matrix (*H*_*tissue*_) by computing the Frobenius norm error (defined as the root square of the sum of squared element-wise differences). These errors were averaged across 100 simulation replicates under four simulation settings, corresponding to *m* = 100, 200, 300, or 400 causal SNPs for the focal disease/trait. We determined that J-PEP attained the lowest average Frobenius norm error for both *H*_*tissue*_ and *H*_*trait*_, with improvements becoming more pronounced as the number of causal SNPs increased (**Fig. 2b** and **Supplementary Table 2**). This indicates that J-PEP’s enhanced predictive performance is not solely driven by the constraint of identifying shared structures between traits and tissues, which is enforced by the third term of the optimization function in Eq. 1, but also benefits from the first two terms of Eq. 1, which ensure that the inferred cluster memberships (*W*) remain closely aligned with the auxiliary trait and tissue data.

Finally, we evaluated the accuracy of Pleiotropic partitioning, Epigenomic partitioning and J-PEP using our new metric (PEPA) and its two constituent metrics (PPA and EPA). The average PEPA achieved by J-PEP increased with larger values of *m*, showing progressively greater and more significant improvements over both Pleiotropic and Epigenomic partitioning as *m* increased (**Fig. 2c** and **Supplementary Table 2)**. At *m* = 200, J-PEP outperformed Pleiotropic and Epigenomic partitioning by 18% (paired *t*-test p = 1.2 × 10^−4^) and 8% (p = 0.04), respectively; at *m* = 400, these gains rose to 19% (p = 2.0 × 10^−10^) and 38% (p = 7.8 × 10^−20^). Among the two constituent methods, Pleiotropic partitioning performed better at predicting SNP-to-tissue associations (*V*_*tissue*_) from SNP-to-trait associations (*V*_*trait*_) (EPA), while Epigenomic partitioning performed better at predicting SNP-to-trait associations (*V*_*trait*_) from SNP-to-tissue associations (*V*_*tissue*_) (PPA); this is consistent with Pleiotropic partitioning (resp. Epigenomic partitioning) yielding lower error when reconstructing the SNP-to-cluster membership matrix (*W*) from SNP-to-trait (resp. SNP-to-tissue) association data, as evaluated in the EPA (resp. PPA) metric—compared to reconstructing *W* from SNP-to-tissue (resp. SNP-to-trait) data, as evaluated in PPA (resp. EPA) (see **Supplementary Note** for a formal derivation of this result).

We performed five secondary analyses. First, we assessed the number of clusters identified by each method at *m* = 400. We observed an average of 2.7 clusters for J-PEP, 2.1 for Pleiotropic partitioning, and 3.9 for Epigenomic partitioning. Second, we performed simulations with varying amounts of uncorrelated structure (**Methods**). We determined that the performance improvement of J-PEP increased in simulations with additional uncorrelated structure, but dissipated in simulations with no uncorrelated structure (**Supplementary Fig. 1**); in the latter case, the information needed to predict SNP-to-trait associations (*V*_*trait*_) or SNP-to-tissue associations (*V*_*tissue*_) can be fully captured by tissue or auxiliary trait data alone. Third, we determined that increasing the number of causal clusters (*K*_causal_) led to reduced prediction accuracy across all methods, though J-PEP consistently achieved the highest PEPA values (**Supplementary Fig. 2a**). Fourth, reducing the maximum number of inferred clusters *K* improved PEPA, suggesting benefits from regularization (**Supplementary Fig. 2b**; also see below). Fifth, we performed simulations in which half of the causal tissues were missing from the epigenomic data, but all relevant auxiliary traits were present (a scenario expected to occur in real data). In these simulations, J-PEP attained greater improvements over Pleiotropic partitioning—outperforming it by 30% on average compared to 19% when all causal tissues were present (**Supplementary Fig. 2c**).

### Application of J-PEP to 165 diseases and complex traits

We applied J-PEP, Pleiotropic partitioning and Epigenomic partitioning to GWAS summary statistics for 165 focal diseases/traits (average of 290K samples and 77 fine-mapped loci; **Supplementary Table 1**), focusing our comparisons on 38 approximately independent diseases/traits for which all three methods identified at least two clusters. Across these 38 traits, J-PEP attained significantly higher (resp. lower) PEPA than Pleiotropic partitioning (genomic block-jackknife *p* < 0.05) for 9 traits (resp. 2 traits) and significantly higher (resp. lower) PEPA than Epigenomic partitioning for 6 traits (resp. 0 traits) (**Fig. 3a** and **Supplementary Table 3**). J-PEP attained the most significant improvements for immune-related traits (e.g., blood cell counts; **Fig. 3a**) and metabolic traits (e.g., total cholesterol; **Supplementary Table 3**). In contrast, Pleiotropic partitioning outperformed J-PEP for one brain-related trait (fractional anisotropy; **Supplementary Table 3**) and one lung-related trait (FEV1) (**Supplementary Table 3**); in each case, J-PEP’s inferred cluster tissue profiles (*H*_*tissue*_) lacked associations with brain and lung annotations, respectively, despite significant enrichment of these and other tissues in the corresponding SNP-to-tissue association matrix (*V*_*tissue*_) matrices.

**Fig. 3.**
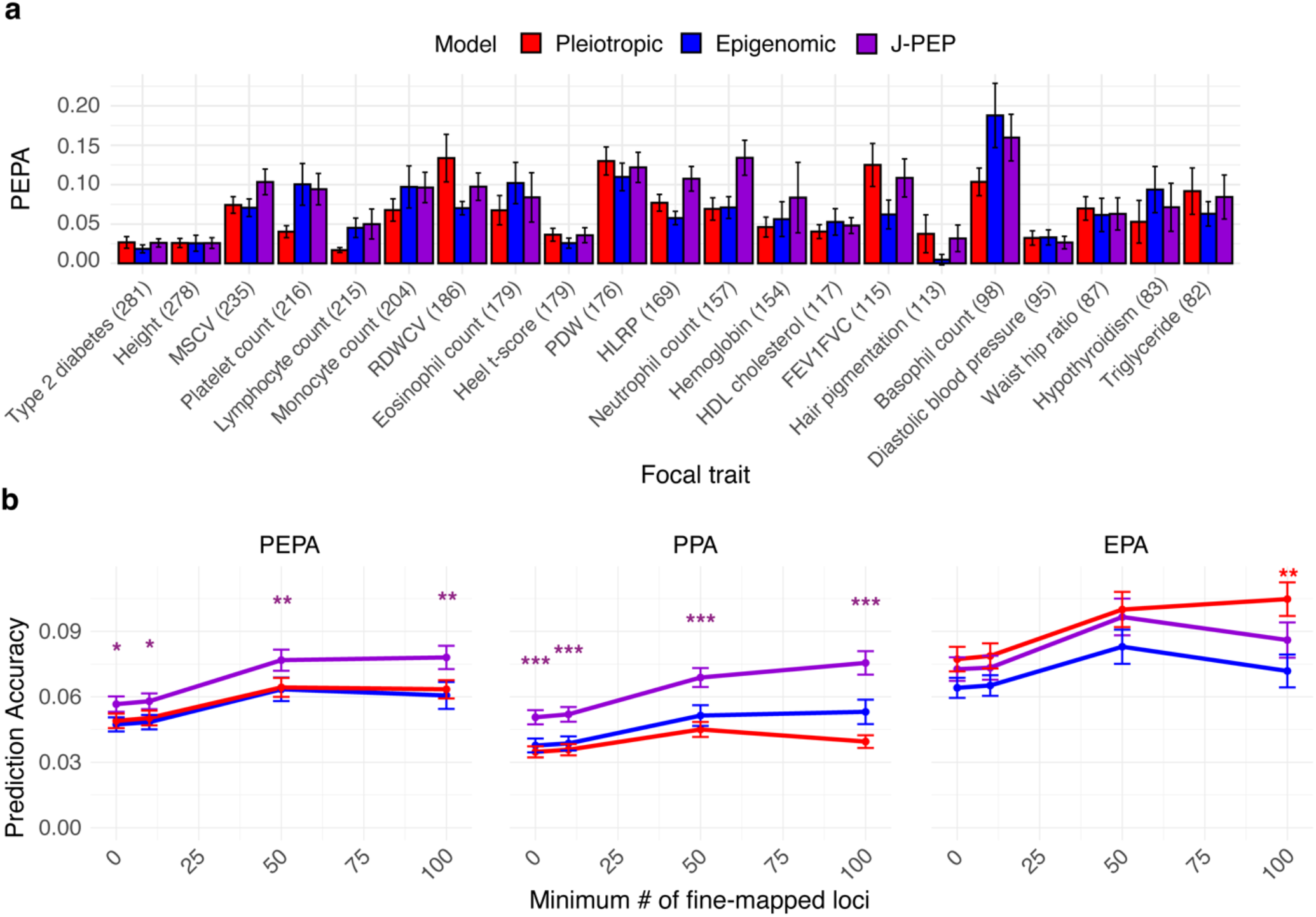
J-PEP outperforms Pleiotropic partitioning and Epigenomic partitioning in analyses of real diseases/traits. (a) PEPA prediction accuracy metric for 20 focal diseases/traits with the largest number of fine-mapped loci (denoted in parentheses). Error bars denote standard errors (via genomic block-jackknife). (b) PEPA, PPA and EPA prediction accuracy metrics averaged across 38, 33, 24, or 16 focal traits (of 38 total) with at least 0, 10, 50 or 100 fine-mapped loci, respectively. Error bars denote standard errors (via genomic block-jackknife). *: p < 0.05, **: p < 0.01, ***: p < 0.001 (vs. each other method). Numerical results for all 165 focal diseases/traits are reported in **Supplementary Table 3**. MSCV: Mean sphered corpuscular volume; RDWCV: Red cell distribution width - coefficient of variation; PDW: Platelet distribution width; HLRP: High light reticulocyte proportion; FEV1FVC: Forced expiratory volume in 1 second/forced vital capacity.

Averaging across the 38 focal diseases/traits, J-PEP attained 16% higher PEPA than Pleiotropic partitioning (*p* = 2.4 × 10^−2^; *p* from inverse-variance weighting [*p*_w_] = 1.2 × 10^−3^; genomic-block jackknife on the average across 38 traits; **Methods**) and 22% higher PEPA than Epigenomic partitioning (*p* = 2.6 × 10^−4^; *p*_w_ = 3.2 × 10^−5^) (**Fig. 3b** and **Supplementary Table 3**). These improvements are consistent with our primary simulations, suggesting that real disease/trait data harbors uncorrelated structure, i.e. structure within *V*_*tissue*_ (resp. *V*_*trait*_) that is not correlated to any structure in *V*_*trait*_ (resp. *V*_*tissue*_). Notably, J-PEP’s performance advantage grew more pronounced in analyses of well-powered traits with more fine-mapped loci, also consistent with simulation results. For example, J-PEP outperformed Pleiotropic partitioning by 27% (*p* = 1.6 × 10^−3^; *p*_w_ = 2.0 × 10^−3^) and Epigenomic partitioning by 30% (*p* = 1.4 × 10^−8^; *p*_w_ = 3.1 × 10^−8^) across 16 focal traits with at least 100 fine-mapped loci (**Fig. 3b** and **Supplementary Table 3**). Among the two constituent methods, Pleiotropic partitioning performed better at predicting SNP-to-tissue associations (*V*_*tissue*_) from SNP-to-trait associations (*V*_*trait*_) (EPA), while Epigenomic partitioning performed better at predicting SNP-to-trait associations (*V*_*trait*_) from SNP-to-tissue associations (*V*_*tissue*_) (PPA), consistent with our simulations. As expected, a given constituent method tends to perform well when J-PEP significantly outperforms the other constituent method (**Supplementary Fig. 3**). These results demonstrate that J-PEP consistently improves tissue–trait prediction accuracy across a broad set of diseases/traits, particularly in analyses of well-powered traits.

We performed eight secondary analyses. First, we examined the number of clusters identified by each method (averaged across 38 focal traits). J-PEP identified fewer clusters on average (2.6) than Pleiotropic partitioning (3.3) and Epigenomic partitioning (3.8), consistent with J-PEP’s objective of suppressing uncorrelated structure (results for each trait are reported in **Supplementary Table 3**). These results were consistent with simulations with the exception of Pleiotropic partitioning, which identified more clusters in real data (3.3) than in simulations (2.1); we hypothesize that this discrepancy may be due to the greater number of auxiliary traits in real data (58 on average) compared to simulations (20), as increasing the number of auxiliary traits in simulations would have required substantially more computational resources. Second, we investigated whether the number of clusters is impacted by GWAS power. For J-PEP, the number of inferred clusters was positively correlated with the number of fine-mapped loci (Pearson *r* = 0.34, p = 1.5 × 10^−2^), indicating that well-powered studies allow finer resolution of underlying processes (**Supplementary Fig. 4**). This relationship was slightly weaker for Pleiotropic partitioning (*r* = 0.27, p = 0.06) and absent for Epigenomic partitioning (*r* = –0.13, p = 0.37). Third, we evaluated how confidently the clusters differentiated each SNP by computing the maximum cluster proportion for each SNP (averaged across focal trait-SNP pairs). All three methods achieved strong differentiation, with mean values of 0.82 (J-PEP), 0.88 (Pleiotropic partitioning), and 0.81 (Epigenomic partitioning) (**Supplementary Fig. 5**); SNPs within the same fine-mapped locus were typically assigned to the same cluster, with concordance rates of 83% (J-PEP), 89% (Pleiotropic partitioning), and 72% (Epigenomic partitioning), based on the maximum summed assignment proportion across clusters, averaged across focal trait-locus pairs. Fourth, we analyzed J-PEP’s aggregated tissue profiles, summing scores across clusters (rows of *H*_*tissue*_) for each focal disease/trait (**Supplementary Fig. 6**, left panel). We detected strong associations to biologically relevant tissues—such as immune cells for blood cell traits, heart for diastolic blood pressure, kidney for kidney volume, and liver for total cholesterol. Because these scores are summed across clusters—each capturing distinct tissue profiles—additional associations also emerged. For example, T2D exhibited associations with seven distinct tissue categories, consistent with diverse contributing processes^23–25^, and neutrophil count exhibited associations to both central nervous system and liver tissues beyond immune cells (these two focal traits are explored further below). Fifth, we sought to validate trait–tissue coherence (beyond PEPA) using an alternative metric, projection correlation, that quantifies the agreement between SNP-level trait and tissue projections across clusters (averaged across 38 focal traits) (**Methods**). J-PEP achieved the highest average projection correlation (*r* = 0.11), outperforming Pleiotropic partitioning (*r* = 0.09; genomic block-jackknife for the difference with J-PEP: p = 0.17) and Epigenomic partitioning (*r* = 0.06; p = 2.8 × 10^−4^) (results for each trait are reported in **Supplementary Table 4**). Sixth, we tested whether J-PEP performance was sensitive to the maximum number of clusters (*K*) by reducing *K* from 15 to 6. This reduced the number of clusters detected by each method by 7-20% but improved the PEPA of each method by 17-23%, with J-PEP maintaining 13% higher PEPA than Pleiotropic partitioning (p = 2.6 × 10^−2^) and 24% higher PEPA than Epigenomic partitioning (p = 6.8 × 10^−5^) (**Supplementary Fig. 7**), consistent with simulations. After considering the tradeoff between the biological information in additional clusters vs. the improvement in PEPA when reducing the maximum number of clusters, we opted to retain the default value of *K* = 15. Seventh, we examined whether tissues with more annotated chromatin features exhibited higher tissue-to-cluster profile scores (*H*_*tissue*_). Indeed, average tissue-to-cluster profile scores across the 38 traits were positively correlated with the number of epigenomic annotations per tissue for each method, with *r* = 0.45 (J-PEP), *r* = 0.69 (Pleiotropic partitioning), and *r* = 0.48 (Epigenomic partitioning) (**Supplementary Table 5**). This implies that tissues with denser annotations are more frequently implicated by all three methods. (We note that Pleiotropic partitioning as defined in this study does not use tissue data for clustering but relies on *V*_*tissue*_ and the inferred *W* to derive *H*_*tissue*_, and is therefore still impacted by annotation density.) The bias due to annotation density should be taken into account when interpreting tissue-to-cluster profiles (see Discussion). Finally, we assessed the running time of J-PEP, as well as Pleiotropic partitioning and Epigenomic partitioning. We determined that all methods had low running times, ranging from 36 to 449 seconds per trait on average for Epigenomic partitioning and Pleiotropic partitioning, respectively (**Supplementary Fig. 8**). All reported times correspond to ten repeated optimization runs per trait-method pair, as required by the default settings of each model (**Methods**).

**Fig. 4.**
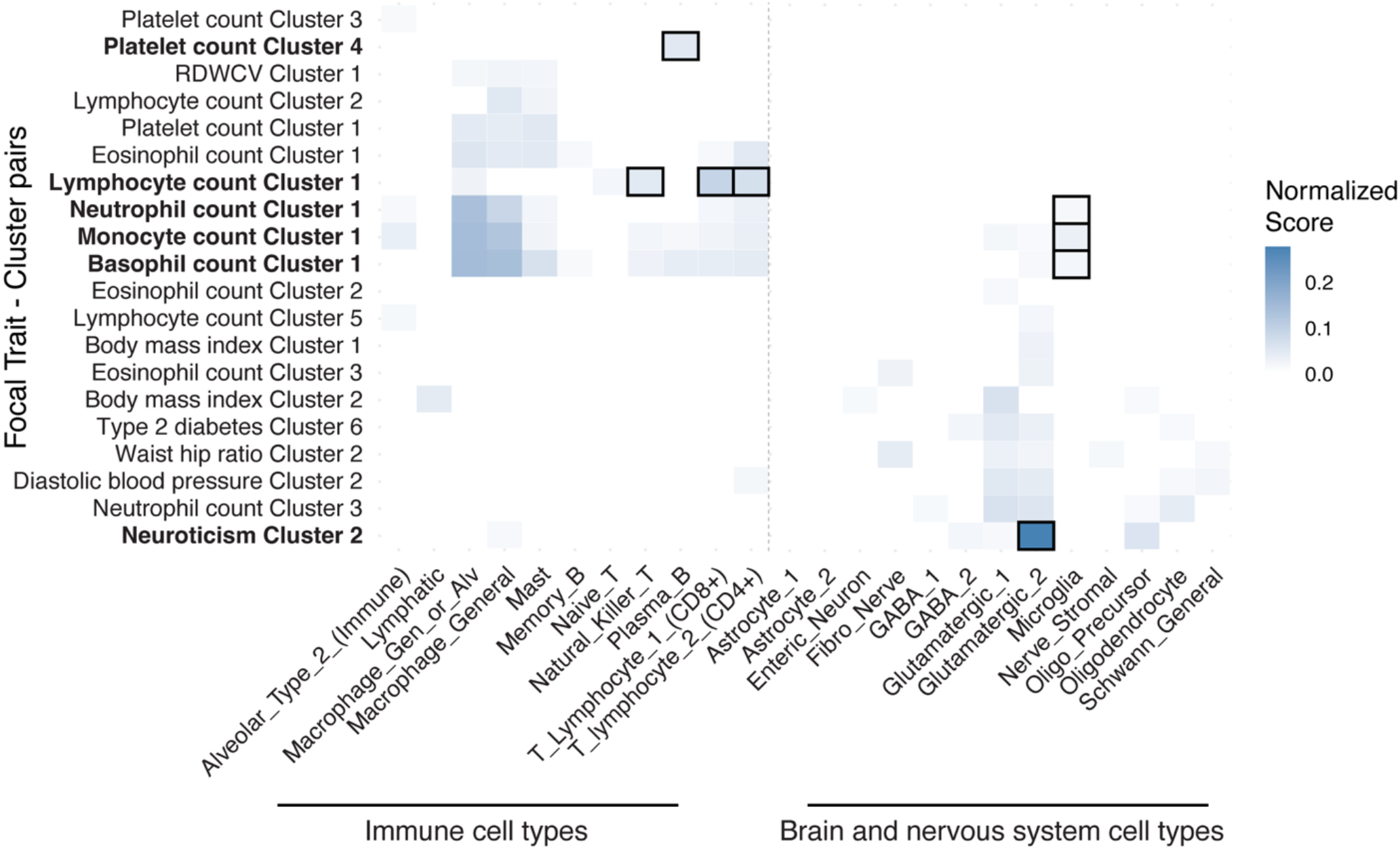
Single-cell data improves resolution and specificity of cell-type associations. We report normalized cell-type scores for each of 20 selected focal trait-cluster pairs (selected as described below; rows) across 24 single-cell derived immune cell types (left) and brain/nervous system cell types (right). Normalized scores are defined as the proportion of each cluster assigned to a given cell type, normalized across all cell types for that cluster. We selected focal trait-pairs with at least one normalized score >1% and sorted them by identifying the tissue category (immune or brain) with the highest aggregate score for each pair, then sorting first by tissue category and second by the magnitude of the maximum aggregate score within that category. Black borders denote focal trait-cluster-cell-type triplets discussed in the main text, with corresponding focal trait-cluster pairs in bold font. Numerical results for 38 diseases/traits are reported in **Supplementary Table 6**. RDWCV: Red cell distribution width - coefficient of variation.

**Fig. 5.**
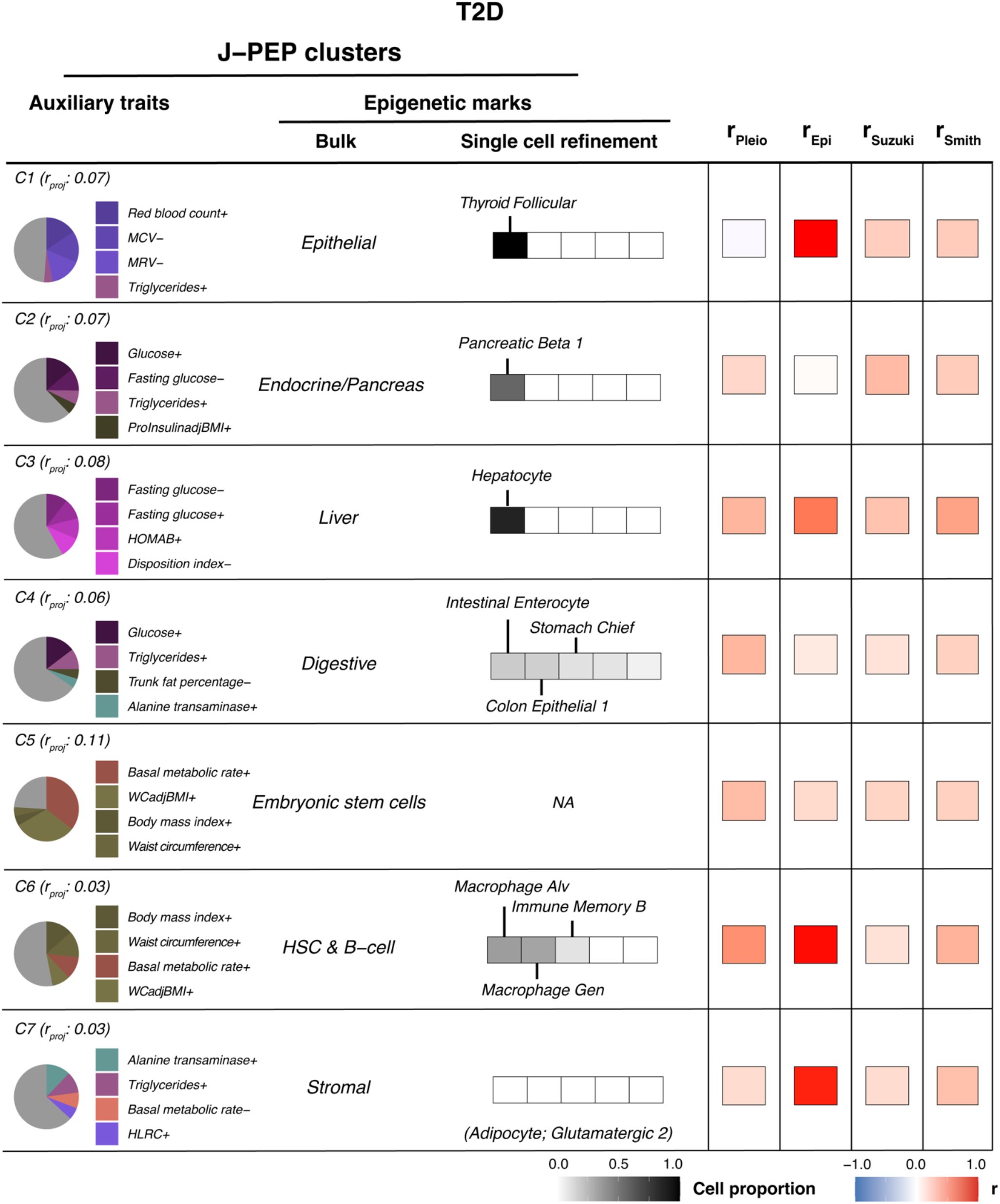
Comparison of T2D clusters identified by J-PEP with other clustering methods. For each T2D cluster identified by J-PEP, the pie chart (left) shows the pleiotropic auxiliary trait profile (top 4 traits), and the bulk and single-cell columns show the associated epigenomic tissue and cell type profiles, respectively. The “+” (resp. “−”) symbol appended to auxiliary trait names denotes pleiotropic associations that are concordant (resp. discordant) with the focal trait (**Methods**). Single-cell refinement results show the top 5 contributing cell types within the most enriched bulk tissue category, shaded by relative cell proportion. NA indicates that the single-cell atlas that we employed lacks cell types from the implicated bulk tissue. Cell types that do not correspond to a refinement of the implicated bulk tissue are denoted in parentheses. Cluster labels indicate the projection correlation (*r*_*proj*_), quantifying internal concordance between trait and tissue profiles (**Methods**); clusters are ordered by the proportion of total variance they explain jointly across traits and tissues (**Methods**). The heatmap shows the maximum correlation between each J-PEP cluster and clusters obtained from Pleiotropic partitioning, Epigenomic partitioning, or two previous T2D clustering studies (Smith et al.^25^ and Suzuki et al.^24^). See Data Availability for numerical results. MCV: Mean corpuscular volume; MRV: Mean reticulocyte volume; ProInsulinadjBMI: Proinsulin adjusted for body mass index; HOMAB: Homeostasis model assessment of β-cell function; WCadjBMI: Waist circumference adjusted for body mass index; HLRC: High light reticulocyte count.

**Fig. 6.**
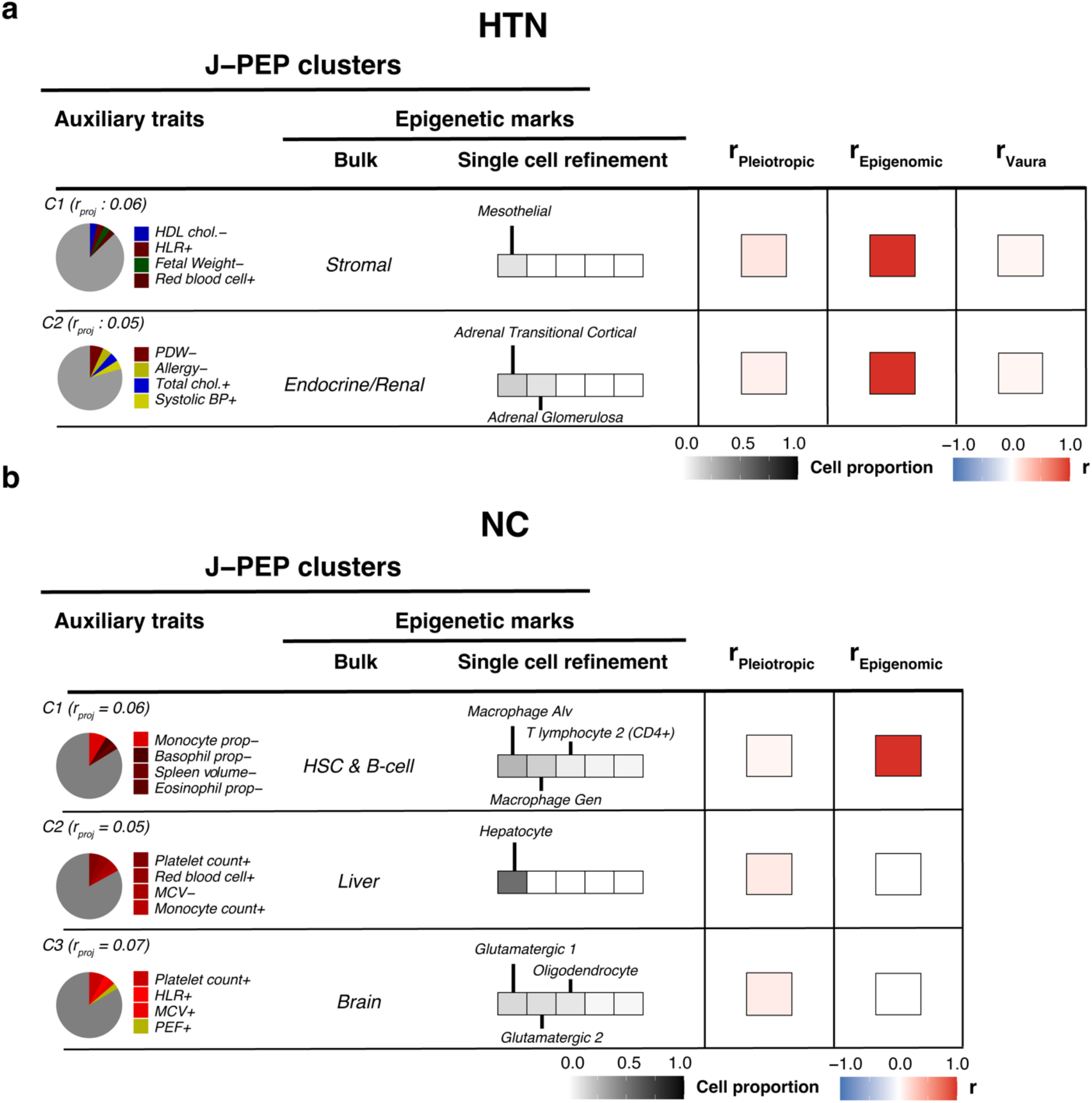
Comparison of HTN and NC clusters identified by J-PEP with other clustering methods. For each hypertension (HTN) cluster (a) and neutrophil count (NC) cluster (b) identified by J-PEP, the pie chart (left) shows the pleiotropic auxiliary trait profile (top 4 traits), and the bulk and single-cell columns show the associated epigenomic tissue and cell type profiles, respectively. Single-cell refinement results show the top 5 contributing cell types within the most enriched bulk tissue category, shaded by relative cell proportion. The “+” (resp. “−”) symbol appended to auxiliary trait names denotes pleiotropic associations that are concordant (resp. discordant) with the focal trait (**Methods**). Cluster labels indicate the projection correlation (*r*_*proj*_), quantifying internal consistency between tissue and trait profiles (**Methods**); clusters are ordered by the proportion of total variance they explain jointly across traits and tissues (**Methods**). The heatmap shows the maximum correlation between each J-PEP cluster and clusters obtained from Pleiotropic partitioning or Epigenomic partitioning (or Vaura et al.^57^ for HTN). See Data Availability for numerical results. HLR: High light reticulocyte count; PDW: Platelet distribution width; BP: Blood pressure; MCV: Mean corpuscular volume; PEF: Peak expiratory flow.

### Single-cell accessibility profiles enhance cell-type resolution and specificity

We sought to assess whether single-cell chromatin accessibility profiles could help interpret the J-PEP clusters for 38 focal disease/trait clusters identified by J-PEP using bulk epigenomic data. We analyzed a single-cell atlas of chromatin accessibility comprising 615,998 cells across 111 fine-grained adult human cell types^42^. We derived cell-type-to-cluster profile matrices (*H*_*cell − type*_) by solving *V*_*cell − type*_ = *W* × *H*_*cell − type*_ using a non-negative least squares approach with multiplicative update rules analogous to those used in bNMF, where *W* denotes the SNP-to-cluster membership matrix inferred by J-PEP using bulk data, and *V*_*cell − type*_ denotes the SNP-to-cell-type association matrix obtained using the EM algorithm on scATAC-seq data across the 111 cell types (analogous to *V*_*tissue*_ as defined above; **Supplementary Table 6**; **Methods**). We compared *H*_*cell − type*_ to *H*_*tissue*_ by collapsing both to 24 unified tissue categories (**Supplementary Table 7**); we observed high concordance (mean *r* = 0.55; **Supplementary Fig. 6**), confirming high concordance between the two modalities. We did not include direct application of J-PEP to single-cell data in our primary analyses; due to increased sparsity, cell-type complexity, and technical variability of single-cell assays, this led to lower prediction accuracy than application of J-PEP to bulk data (although J-PEP still substantially outperformed Pleiotropic partitioning and Epigenomic partitioning in direct application to single-cell data; **Supplementary Fig. 7**).

We evaluated whether scATAC-seq data could improve the resolution and specificity of cell-type associations. Restricting to the 11 of 24 unified tissue categories with at least four annotated cell types, we determined that focal trait-cluster pairs positively associated to these tissue categories were associated to only 21% of the corresponding cell-types on average (**Fig. 4, Supplementary Fig. 9** and **Supplementary Table 6**), indicating a high degree of specificity. For example, in the immune category, cluster 1 of lymphocyte count was predominantly associated with T-cell subtypes, whereas cluster 4 of platelet count was exclusively linked to plasma B cells. In the brain and nervous system category, cluster 2 of neuroticism was specifically associated with a glutamatergic neuron subtype—excitatory cells critical for brain signaling— whereas cluster 1 of each white blood cell trait (neutrophil count, monocyte count and basophil count) showed distinct associations with microglia, the brain’s resident immune cells. In the skin category, which comprises four annotated cell types, cluster 1 of hair pigmentation was uniquely associated with melanocytes, the pigment-producing cells of the epidermis (**Supplementary Fig. 9**). These findings demonstrate that single-cell epigenomic data can resolve the cellular basis of trait–tissue associations, enhancing the interpretability of J-PEP-derived clusters.

### J-PEP identifies non-canonical axes of type 2 diabetes pathogenesis

Type 2 diabetes (T2D) is a multifactorial disorder involving heterogeneous pathological mechanisms including pancreatic β-cell dysfunction, impaired insulin production by excess hepatic fat accumulation and obesity-related insulin resistance^23–25,46,47^. Prior efforts to partition T2D loci—often via pleiotropic partitioning—have consistently identified clusters reflecting these distinct processes^23–25,27,30,31,48^. However, these approaches have often lacked resolution into the specific cellular contexts in which they occur, a limitation that J-PEP is designed to address. To enable direct comparison with previous work, all results in this section are based on the 109 auxiliary traits from the most comprehensive pleiotropic partitioning study to date^25^ (**Supplementary Table 8**), which employed a method closely aligned with Pleiotropic partitioning as defined in the current study. Results based on the default set of 82 auxiliary traits (see *Overview of methods*) are provided in the **Supplementary Note** (PEPA values in **Figure 3A**).

We applied J-PEP to T2D summary statistics from ref. ^24^ (428,452 cases, 2,107,149 controls). J-PEP identified seven clusters, capturing both established and underexplored biological processes (**Fig. 5**). With respect to the PEPA metric, J-PEP (PEPA = 0.03) did not significantly differ from Epigenomic partitioning (PEPA = 0.04, p = 0.28; 5 clusters), and was outperformed by Pleiotropic partitioning (PEPA = 0.06, p = 5.1 × 10^−4^; 5 clusters) (**Supplementary Table 3**), consistent with the tradeoff between the biological information in additional clusters vs. optimizing PEPA (**Supplementary Fig. 7**). We detail J-PEP’s results below, as its clusters integrate elements from both pleiotropic and epigenomic partitions, offering a broad perspective on disease processes.

For each J-PEP cluster, we computed SNP-level correlations between each cluster produced by a different method (defined as the correlation between the corresponding columns of the respective SNP-to-cluster membership matrices *W*) and assessed the maximum correlation. For clusters 2,4, and 5, SNP-level correlations were strongest between J-PEP and pleiotropy-based methods (including Smith et al.^25^ and Suzuki et al.^24^), yet overall correlations remained modest (mean max *r* = 0.34; **Fig. 5**); for clusters 1, 3, 6, and 7, correlations were strongest between J-PEP and Epigenomic partitioning, with high correlations (mean max *r* = 0.96; **Fig. 5**), underscoring the informativeness of tissue data.

Clusters 2–4 align with canonical T2D pathways^23–25,27,30,31,48^ (**Fig. 5**). Cluster 2 is associated with insulin secretion traits and endocrine/pancreatic tissues and is enriched for GO terms related to insulin signaling and glucose regulation (**Supplementary Table 9** and **Methods**). Notably, single-cell data refined this association to pancreatic islet β-cells, supporting a β-cell dysfunction mechanism contributing to insulin deficiency^23,24,49,50^. Cluster 3 exhibits a consistent liver signature across bulk and single-cell profiles with the single-cell association refined to hepatocytes—the main liver cell type and the only one represented in the single-cell data— and GO enrichment for cholesterol efflux and triglyceride metabolism, indicative of hepatic lipid dysregulation contributing to insulin resistance^23–25,27^. Cluster 4 is associated with liver enzyme and adiposity traits and maps to digestive tissues. Single-cell data refined this digestive association to multiple gastrointestinal cell types and GO term enrichment for hydrogen peroxide catabolism points to hepatic oxidative stress linked to lipid overload and steatosis—a known mechanism of insulin resistance^7^.

Clusters 1 and 5–7 reflect non-canonical components of T2D biology (**Fig. 5**). Cluster 1 is associated with red blood cell traits and epithelial tissues. Single-cell data refined this association to thyroid follicular cells, and GO enrichment revealed epithelial-to-mesenchymal transition and oxidative stress, suggesting a stress-related process potentially affecting insulin signaling^51^. Cluster 5 lacks a clear pathological interpretation but is enriched for chromatin organization and associated with embryonic stem cells. Refinement using single-cell data was not feasible, as the single-cell atlas that we employed lacks embryonic cell types. This cluster may reflect early developmental or transcriptional regulatory influences on T2D risk. Cluster 6 is associated with obesity-related traits and immune tissues. Single-cell data refined this association to macrophages, with GO enrichment for inflammatory signaling and thermogenesis, suggesting a role for immune-metabolic regulation of energy balance in T2D^52–54^. Finally, Cluster 7 is characterized by associations with liver enzymes and triglycerides, GO enrichment for lipid homeostasis pathways, and a stromal bulk tissue profile refined by single-cell data to a mix of adipocyte and neuronal cell types, suggesting a metabolic mechanism.

### J-PEP identifies stromal and endocrine axes of hypertension pathogenesis

Hypertension (HTN) is a highly polygenic disease with contributions from multiple biological pathways^55,56^. Prior clustering efforts have often focused on shared metabolic comorbidities such as obesity, lipid levels, or T2D, providing limited insight into tissue-specific regulatory contributions to blood pressure control^57,58^.

We applied J-PEP to HTN summary statistics from ref. ^41^ (3,174 cases, 455,380 controls), using 68 auxiliary traits (**Supplementary Table 8**). J-PEP identified two clusters described in detail below (**Fig. 6a**). With respect to the PEPA metric, J-PEP (PEPA = 0.07) outperformed both Pleiotropic partitioning (PEPA = 0.04, p = 0.07; 3 clusters) and Epigenomic partitioning (PEPA = 0.03, p = 2.1 × 10^−3^; 5 clusters) (**Fig. 3a, Supplementary Table 3**).

Cluster 1 is associated with bulk stromal tissue, refined by single-cell data to mesothelial cells, with GO enrichment for Wnt signaling and cytoskeletal remodeling (**Supplementary Table 9)**, suggesting mesothelial-driven extracellular matrix (ECM) remodeling and vascular stiffening, a known contributor to elevated vascular resistance in hypertension^59^. While ECM remodeling is a recognized mechanism in hypertension pathophysiology^60,61^, the specific role of mesothelial cells in this process remains speculative^62^. Cluster 2 is associated with endocrine bulk tissue, refined by single-cell data to adrenal cortex cells, with GO terms highlighting neutrophil-mediated immunity, indicating a potential feedback between aldosterone signaling and local immune activity^63^. Although aldosterone is known to modulate various immune pathways involved in inflammation—and immune-targeted therapies have shown benefit in managing hypertension— the precise mechanisms underlying its crosstalk with the immune system remain unknown^64^. For both cluster 1 and cluster 2, SNP-level correlations were strongest between J-PEP and Epigenomic partitioning (**Fig. 6a**), highlighting the benefit of incorporating epigenomic data. Compared to the four metabolically themed clusters identified by Vaura et al.^57^, who applied a method similar to Pleiotropic partitioning to FinnGen data^65^—comprising obesity, two lipid-related, and one stature-associated clusters— J-PEP produced a distinct set of clusters with minimal overlap—likely reflecting its integrative design that incorporates epigenomic information.

### J-PEP identifies immune, hepatic, and neuroinflammatory axes of neutrophil count

Neutrophil count (NC) is a key hematological trait central to immune and inflammatory processes^58,59^, yet to our knowledge it has not previously been analyzed using clustering methods that integrate either pleiotropic or epigenomic data. We applied J-PEP to NC summary statistics from ref. ^66^ (563,085 samples), using 83 auxiliary traits (**Supplementary Table 8**). J-PEP identified three clusters, reflecting immune, hepatic, and neuroinflammatory axes (**Fig. 6b**). With respect to the PEPA metric, J-PEP (PEPA = 0.14) significantly outperformed Pleiotropic partitioning (PEPA = 0.05, p = 3.8 × 10−9; 5 clusters) and Epigenomic partitioning (PEPA = 0.06, p = 6.4 × 10−6; 3 clusters) (**Fig. 3a, Supplementary Table 3**).

Cluster 1 is associated with bulk hematopoietic stem and progenitor cells, refined by single-cell data to alveolar and general macrophages, with GO enrichment for chemotaxis, immune activation, and antimicrobial responses (**Supplementary Table 9**), highlighting canonical immune regulation of neutrophils^67^. Cluster 2 exhibits a consistent liver signature across bulk and single-cell profiles (refined to hepatocytes in single-cell data), and is linked to platelet-related traits, suggesting a speculative hepatic-inflammatory connection to neutrophil regulation, consistent with neutrophil activation in metabolic disorders^68^. Cluster 3 exhibits a bulk brain profile refined by single-cell data to glutamatergic neurons and oligodendrocytes, along with associations to hematocrit and platelet count, pointing to emerging but unconfirmed links between neural regulation and immune tone^69,70^. SNP-level correlations indicate that Cluster 1 aligns closely with Epigenomic partitioning, whereas Clusters 2 and 3 are only weakly recovered by Pleiotropic partitioning—despite their most distinctive features (compared to Cluster 1 and to each other) being reflected in their tissue and cell-type profiles, which notably do not implicate immune-related components.

## Discussion

We have developed J-PEP, a method that integrates pleiotropic and epigenomic information to partition disease loci into biologically interpretable clusters. By leveraging shared structure across auxiliary traits and tissues, J-PEP improves the accuracy of predicting SNP-to-trait and SNP-to-tissue associations, which we quantify using our new PEPA metric. These gains are most pronounced for focal diseases/traits whose underlying biology is well-captured by available epigenomic datasets, such as immune- and liver-related traits. In our applications to GWAS summary statistics for a broad set of diseases and traits, J-PEP recapitulates canonical disease pathways, e.g. endocrine/pancreatic dysfunction and hepatic-driven insulin resistance for type 2 diabetes^23–25^, but also reveals underexplored biological processes, e.g. immune-metabolic and developmental components for type 2 diabetes. Notably, incorporating single-cell epigenomic profiles refines tissue associations to cell-type resolution, e.g. resolving an endocrine/pancreatic cluster for type 2 diabetes to pancreatic β-cells, and resolving an endocrine/renal cluster for hypertension to specific adrenal cortex cell types.

J-PEP shares conceptual goals with several recent approaches aimed at disentangling shared genetic architecture across complex traits. DeGAs^17^ applies truncated singular value decomposition (tSVD) to GWAS summary statistics to extract latent components of genetic variation. While scalable and effective for broad surveys, it lacks structured priors to promote sparsity, usually requiring post-hoc enrichment analyses for interpretation. FactorGo^26^ addresses this by modeling uncertainty in GWAS effect estimates within a variational Bayesian framework and promoting sparsity through automatic relevance determination. However, it operates on LD-pruned data, which limits compatibility with regulatory annotations that do not depend on LD structure, such as epigenomic marks. The V2G2P framework^38^ takes a more mechanistic route, linking GWAS variants to genes through enhancer maps and to regulatory programs derived from single-cell perturbation screens. This strategy enables interpretable, cell-type-specific disease insights, but it requires substantial experimental resources and is currently limited to well-characterized cellular systems, limiting its scalability.

Our results have several downstream implications. From a translational perspective, J-PEP provides a scalable means to distill complex polygenic signals into a small number of interpretable axes of biological activity. This may aid in prioritizing genes, tissues, and pathways for functional follow-up and therapeutic development^4,71,72^. Also, by uncovering shared axes across diseases/traits, J-PEP may help identify opportunities for drug repurposing—for example, targeting pathways implicated in both type 2 diabetes and cardiovascular disease^73,74^. In the context of precision medicine, clusters inferred by J-PEP could aid the construction of partitioned polygenic risk scores, previously shown to support disease subtyping^24,25^. Finally, our PEPA metric provides a principled framework for benchmarking future methods that integrate pleiotropic and epigenomic data.

We note several limitations of our work. First, while J-PEP captures trait–tissue correlations, it does not explicitly infer causality between them; nonetheless, clusters derived from joint partitioning are less prone to spurious associations compared to single-modality approaches, as they better reflect disease-relevant biological context. Second, the algorithm can be computationally demanding for large-scale datasets, particularly when incorporating high-dimensional modalities like single-cell multi-omics; however, the method is computationally tractable (**Supplementary Fig. 8)**, and efficiency can be further enhanced by restricting inputs to critical tissues or a curated set of auxiliary traits. Third, J-PEP assumes a linear shared structure between modalities, which may overlook non-linear interactions between pleiotropy and epigenomic data; however, the linearity assumption is likely to be a reasonable approximation given the largely additive nature of genetic effects^22,75,76^. Fourth, the requirement for concordant clustering across traits and tissues may suppress modality-specific associations (e.g., clusters detectable only through pleiotropy or epigenomics alone); this constraint also underlies the definition of the PEPA metric. However, this restriction likely improves robustness by focusing on clusters supported by multiple lines of evidence. Fifth, although J-PEP outperforms single-modality methods, overall PEPA scores remain modest, reflecting inherent biological complexity, measurement noise, and incomplete regulatory and phenotypic coverage—gaps that are likely to narrow as new genetic and epigenomic resources continue to emerge^4,77–80^. Sixth, our criterion for selecting auxiliary traits (rg < 0.5 with the focal trait) aims to ensure independence from the focal trait but does not constrain genetic correlation among auxiliary traits themselves. This may allow residual pleiotropic structure among auxiliary traits to influence results. Nonetheless, for T2D, we observe that results remain highly consistent across largely independent auxiliary trait sets (see **Supplementary Note**). Finally, in bulk epigenomic data, the results of J-PEP (and other methods) are biased towards tissues with denser annotations; in single-cell epigenomic data, direct application of J-PEP is limited by sparsity and technical variability. Ongoing expansion of tissue-specific and single-cell epigenomic datasets will help mitigate both issues^4,77–80^. Despite these limitations, J-PEP provides a generalizable and interpretable approach for partitioning GWAS loci into biologically distinct clusters.

## Code Availability

We have released open-source software implementing J-PEP and PEPA at https://github.com/kernerg/jpepR.

## Data Availability

SNP-to-cluster membership and cluster-specific trait and tissue profile matrices for Pleiotropic partitioning, Epigenomic partitioning and J-PEP have been made publicly available at https://alkesgroup.broadinstitute.org/J-PEP.

PEPA, PPA and EPA values for all 165 diseases/traits have been made publicly available at https://alkesgroup.broadinstitute.org/J-PEP.

The bulk-based epigenomic data used in this work can be accessed through the EpiMap Repository^36^ (https://compbio.mit.edu/epimap/).

The raw single-cell-based epigenomic data^42^ used in this work can be downloaded at: https://www.ncbi.nlm.nih.gov/geo/query/acc.cgi?acc=GSE184462. Peaks used in this work were downloaded using the following command line: *wget -r --level=1 -np -R index.html* http://catlas.org/catlas_downloads/humantissues/Peaks/.

GWAS summary statistics for the 165 diseases/traits analyzed in this study, as well as the 109 auxiliary traits from ref. ^25^ analyzed in the T2D subsection, are available at https://alkesgroup.broadinstitute.org/J-PEP.

## Acknowledgements

We thank Kushal Dey and all members of Alkes Price’s lab for helpful discussions and data sharing. This research was funded by NIH grants R01 HG006399, U01 HG012009, R01 MH101244, R01 HG013083, R37 MH107649 and T32 HG002295.

## Methods

### J-PEP method: input and output

J-PEP inputs *V*_*trait*_ and *V*_*tissue*_ and outputs *W, H*_*trait*_ and *H*_*tissue*_.

*V*_*trait*_ ∈ *ℝ*^*M*×*A*^ is a SNP-to-auxiliary trait matrix defined for a given focal trait. M is the number of fine-mapped SNPs (PIP >0.01) for the focal trait and *A* is the number of auxiliary traits. The entries of this matrix are defined as:

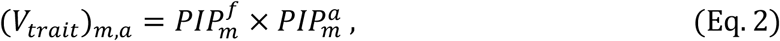

where 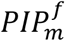 and 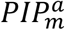 denote the posterior inclusion probabilities (PIPs) for SNP *m* in the focal trait *f* and auxiliary trait *a*, respectively. Because J-PEP relies on non-negative matrix factorization, each auxiliary trait is split into two components: a concordant pleiotropic component (denoted by a “+” sign in figures) and a discordant pleiotropic component (denoted by a “−” sign).

*V*_*tissue*_ ∈ ℝ^*M*×*T*^ is a SNP-to-tissue matrix defined for a given focal trait. *M* is the number of fine-mapped SNPs (PIP >0.01) for the focal trait and *T* is the number of tissues. The entries of this matrix are defined as:

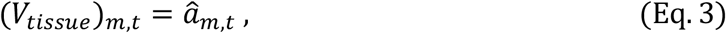

where 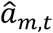 is a normalized score quantifying the strength of association between SNP *m* and tissue *t* derived from tissue-specific epigenomic profiles. We generate this using the E-M algorithm. Details are below (see **EpiMap data set**).

*W* ∈ ℝ^*M*×*K*^ is a SNP-to-cluster membership matrix defined for a given focal trait. *M* is the number of fine-mapped SNPs (PIP >0.01) for the focal trait and *K* is the number of clusters identified by the J-PEP method.

*H*_*trait*_ ∈ ℝ^*K*×*A*^ is a cluster-specific profile matrix for auxiliary traits. *K* is the number of clusters identified by the J-PEP method and *A* is the number of auxiliary traits, as in *V*_*trait*_.

*H*_*tissue*_ ∈ ℝ^*K*×*T*^ is a cluster-specific profile matrix for tissues. *K* is the number of clusters identified by the J-PEP method and *T* is the number of tissues, as in *V*_*tissue*_.

### J-PEP method: objective function

J-PEP performs joint non-negative matrix factorization on *V*_*trait*_ and *V*_*tissue*_, enforcing a shared SNP-to-cluster membership matrix *W*. The objective function is:

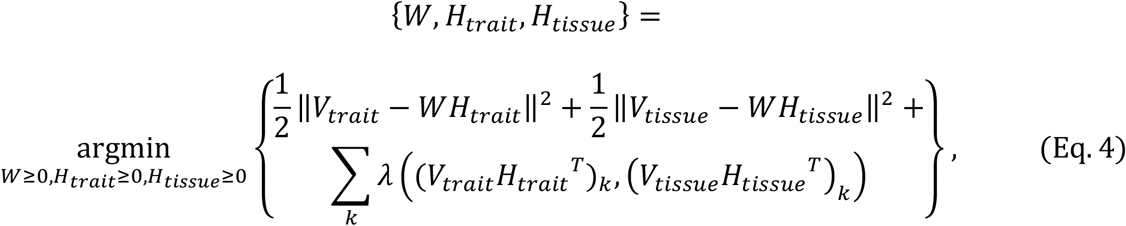

where *k* is taken across the *K* clusters (columns of *V*_*trait*_*H*_*trait*_^*T*^ and *V*_*tissue*_*H*_*tissue*_^*T*^) from the current iteration of the algorithm and the penalty function *λ* promotes alignment between pleiotropic and epigenomic data:

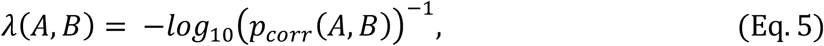

where *p*_*corr*_ indicates the one-sided Pearson’s correlation *p* value for positive correlation.

We note that since *V*_*trait*_*H*_*trait*_^*T*^ and *V*_*tissue*_*H*_*tissue*_^*T*^ approximate the shared cluster membership matrix *W* (and are equal to it when *H*_*trait*_ and *H*_*tissue*_ are orthogonal matrices, though this is almost never the case), the penalty function *λ* encourages clusters where SNP memberships are consistent across trait and tissue data.

We also note that *λ* depends on the number of fine-mapped SNPs, so the method uses a scaling parameter to adjust the strength of the penalty dynamically and can be tuned by the user.

### J-PEP method: optimization

We optimized the J-PEP objective function using an iterative algorithm alternating updates of the shared SNP-to-cluster membership matrix *W*, the trait-specific cluster profile matrix *H*_*trait*_, and the tissue-specific profile matrix *H*_*tissue*_. All matrices were initialized with small positive random values scaled to the input range. Updates were performed using multiplicative update rules derived from the gradient of the objective function, as done in bNMF, incorporating the penalization term *λ*. To maintain numerical stability, a small constant ε = 10^−50^ was added to all matrix approximations.

Convergence was assessed by monitoring relative changes in the penalization weights applied to each cluster. The algorithm terminated when the maximum relative change across iterations fell below a predefined threshold (default: 10^−7^) or when the maximum number of iterations (default: 5,000) was reached. We discarded solutions in which fewer than two non-redundant clusters remained active (defined by row sums of *H*_*trait*_ and *H*_*tissue*_ exceeding a minimal threshold, defined by default to 10^−10^). Additionally, we discarded solutions in which at least one method failed to identify at least two clusters in a minimum of 18 out of the 22 model runs required for computing prediction accuracy metrics (one per chromosome removed; see below). Applying these two criteria retained 38 of the original 55 focal traits from the full set of 165 defined as approximately genetically independent (pairwise rg <0.5).

#### Sparsity constraint

To improve interpretability, J-PEP imposes a sparsity constraint on tissue profiles by assigning each tissue to its dominant cluster during each iteration, whereas auxiliary traits remain free to associate with multiple clusters. For robustness, we perform ten optimization runs with different random seeds and retain the solution with the highest likelihood among those yielding the most consistent number of clusters; in case of ties, we prioritize the run with fewer clusters.

#### Cluster ordering

We ordered clusters based on the proportion of total variance they jointly explained across both trait and tissue association matrices. Specifically, for each cluster *k*, we computed its contribution to the SNP-to-trait matrix *V*_*trait*_ and the SNP-to-tissue matrix *V*_*tissue*_ using rank-1 reconstructions: *V*_*trait,k*_ = *W*_*k*_*H*_*trait*_[*k*..] and *V*_*tissue,k*_ = *W*_*k*_*H*_*tissue*_[*k*.]. The total variance explained by cluster *k* was defined as the sum of squared entries in *V*_*trait,k*_ and *V*_*tissue,k*_, normalized by the total variance of the original matrices. Clusters were then ranked in decreasing order of this joint variance contribution.

### Pleiotropic partitioning method

The Pleiotropic partitioning method factorizes *V*_*trait*_ using bNMF, that is, by solving the optimization problem:

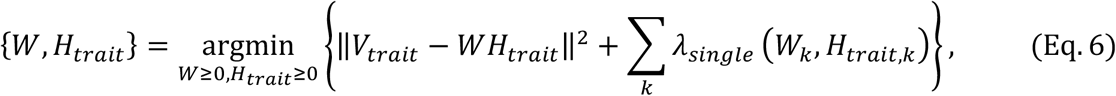

where *W*and *H*_*trait*_ are defined as above, and *λ*_*single*_ is a penalty term that reduces overfitting by pruning irrelevant clusters^32^. The maximum number of clusters *K* is predefined (typically *K* = 15), but the actual number of identified clusters by the partitioning method is often reduced as irrelevant components are penalized to zero. *M* includes all fine-mapped SNPs with PIP values ≥ for the focal trait.

We generate cluster-specific tissue profiles (*H*_*tissue*_) while ensuring that each tissue associates with a single cluster, as follows:

1. We estimate *H*_*tissue*_ from *V*_*tissue*_ and *W* using gradient descent by solving *V*_*tissue*_ = *W* × *H*_*tissue*_, without applying any constraint on *H*_*tissue*_.
2. Clusters with highly similar tissue profiles are merged by averaging their respective cluster memberships. Similarity is assessed using the *simil* function from the R v.4.2 proxy package, with “high similarity” defined as a similarity score > 0.5.
3. We repeat step (1) but now enforce the sparsity condition, as implemented in the J-PEP method.

### Epigenomic partitioning method

The Epigenomic partitioning method factorizes *V*_*tissue*_ using bNMF, that is, by solving the analogous optimization problem to the Pleiotropic partitioning method, using as input *V*_*tissue*_ instead of *V*_*trait*_:

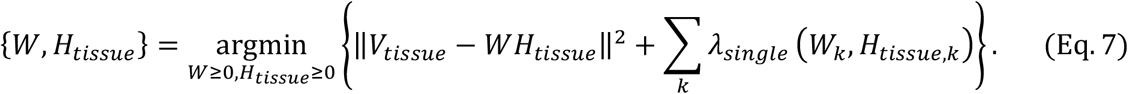

We learn cluster-specific auxiliary trait profiles (*H*_*trait*_) from *V*_*trait*_ and *W* using gradient descent by solving *V*_*trait*_ = *W* × *H*_*trait*_.

### Prediction accuracy metrics

#### PPA

Let *V*_*trait*_ ∈ ℝ^*M*×*A*^ and *V*_*tissue*_ ∈ ℝ^*M*×*T*^ represent the SNP-to-trait and SNP-to-tissue matrices, respectively, where *M* is the number of fine-mapped SNPs (PIP >0.01) for the focal trait, *A* is the number of auxiliary traits, and *T* is the number of tissues. The prediction process aims to estimate *V*_*trait*_ using the SNP-to-cluster membership matrix *W* and the tissue cluster profiles *H*_*tissue*_ and *H*_*trait*_.

Essentially, PPA follows a two-step approach:

1. For a given cluster *k* and a held-out chromosome *c*, we predict the cluster membership 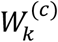 based on *V*_*tissue*_^(*c*)^ and *H*_*tissue*_^−(*c*)^_*k*_, the SNP-to-tissue associations for chromosome *c* and the tissue profiles inferred from the remaining 21 chromosomes, using gradient descent.
2. Then, the predicted SNP-to-trait associations for cluster *k* and chromosome *c*, i.e. 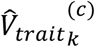, are calculated as:

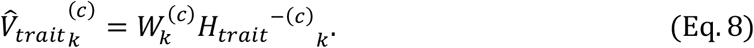

The Pearson correlation between the predicted values 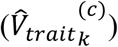 and the true values (*V*_*trait*_^*(c)*^) for each cluster is computed as:

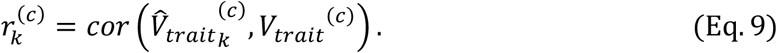

The weighted mean correlation across all clusters in chromosome *c* is computed as:

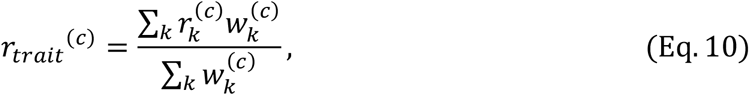

where 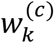 is defined as:

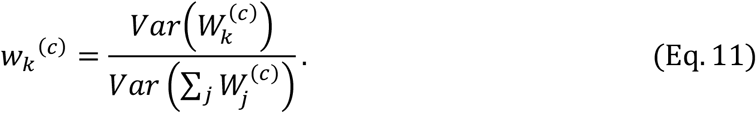

EPA Computation for EPA is analogous to PPA, inverting the roles of trait and tissue data (e.g. *V*_*tissue*_ for *V*_*trait*_ and *V*_*trait*_ for *V*_*tissue*_). The Pearson correlation between the predicted values 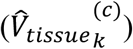 and the true values (*V* ^(1)^) for each cluster is computed as:

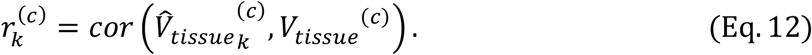

The weighted mean correlation across all clusters in chromosome c is computed as:

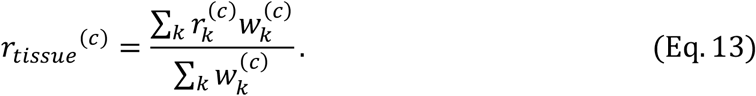

#### PEPA

The overall PEPA score is the sum of the weighted average correlations for both prediction tasks across all chromosomes:

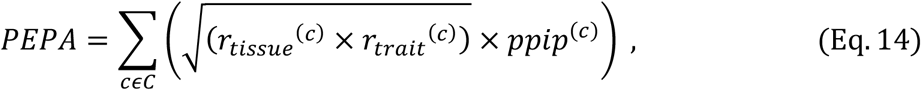

where *ppip*^(*c*)^ denotes the proportion of fine-mapping PIPs on chromosome *c* relative to the total across all held-out chromosomes in the set C, where C indicates the group of chromosomes containing at least five GWAS loci. PEPA was only computed in cases in which |C| > 18.

#### Projection correlation

To validate trait–tissue coherence across identified clusters, we computed a projection correlation metric beyond the PEPA metric, defined as 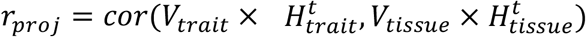.

### Simulation framework

Generating simulated data consists of 8 steps: Step 1: simulating cluster-specific profiles, Step 2: estimating the SNP-to-cluster membership matrix, Step 3: computing SNP-to-trait associations, Step 4: sampling causal SNPs, Step 5: sampling causal effect sizes, Step 6: sampling GWAS summary statistics, Step 7: fine-mapping, Step 8: generating uncorrelated structure.

#### Step 1. Simulating cluster-specific profiles

We initialized cluster-specific trait and tissue profiles as matrices *H*_*trait*_ ∈ ℝ^*A*×*K*^ and *H*_*tissue*_ ∈ ℝ^*T*×*K*^, where *A* is the number of auxiliary traits, *T* the number of tissues, and *K* the number of causal clusters. Each matrix was initialized with standard uniform values, and a subset of traits and tissues was randomly assigned as causal for each cluster (one trait and one tissue per cluster) by retaining non-zero values only in the corresponding columns:

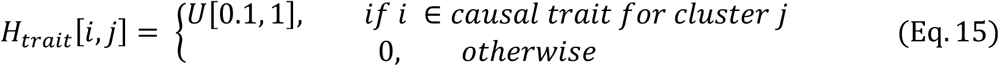

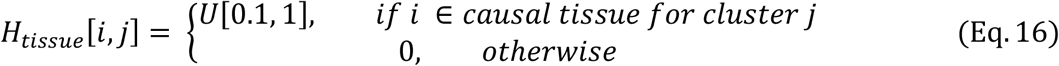

Each row was normalized to sum to one. We used *K* = 5 by default and tested scenarios with *K* = 3 and *K* = 8.

#### Step 2. Estimating SNP-to-cluster membership matrix

The SNP-to-cluster membership *W*, which probabilistically maps SNPs to clusters, was inferred using gradient descent, a SNP-to-tissue matrix from real data analysis and simulated *H*_*tissue*_, by solving the factorization problem: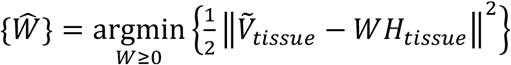. where 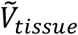 is a SNP-to-tissue matrix constructed by intersecting SNPs from chromosomes 1 and 2 of 1000 Genomes Phase 3^41^ with EpiMap^36^ annotations (see **Epimap data set**). Only SNPs with MAF >0.05 were retained.

#### Step 3. Computing SNP-to-trait associations

The SNP-to-trait matrix *V*_*trait*_ was obtained by matrix multiplication: *W* × *H*_*trait*_.

#### Step 4. Sampling causal SNPs

Causal SNPs for the focal and auxiliary traits were sampled with probability proportional to:

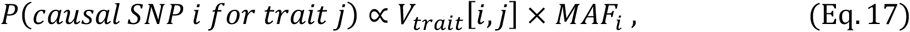

where *MAF*_*i*_ is the MAF of SNP *i*. We assumed the first column of *V*_*trait*_ represents the focal trait.

For auxiliary traits contributing to clusters, we enforced at least 25% overlap in causal SNPs with the focal trait. We varied the number of total causal SNPs *m* ∈ {100; 200; 300; 400} causal SNPs, sampling from different LD blocks on chromosomes 1 and 2.

#### Step 5. Sampling causal effect sizes

Effect sizes were sampled from a standard multivariate normal distribution: *β* ~ *N*(0, *I*_*M*_), where *I*_*M*_ is the identity matrix of dimension equal to the number of causal SNPs. Each trait’s total SNP-heritability 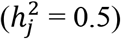 was distributed equally across its causal SNPs by rescaling effect sizes such that their squared sum matched 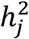.

#### Step 6. Simulating GWAS summary statistics

We then simulated summary statistics using the RSS likelihood^81^, 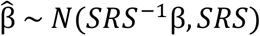, where *β* is the vector of true causal effect sizes, *R* is the LD matrix from chromosomes 1 and 2 of the 1000 Genomes Project Phase 3, *S* is a diagonal matrix with elements 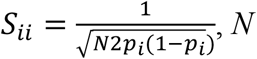 is the sample size and *p* the minor allele frequency of SNP *i*. We used a regularized LD matrix, *R* × (1 − *λ*) + *λI*, where *λ* = 02001.

#### Step 7. Fine-mapping

We performed single causal variant fine-mapping^39,40^ on all simulated GWAS summary statistics as in ref. ^20^. We note that multiple causal variant fine-mapping is not recommended when in-sample LD is not available^82^.

#### Step 8. Generating uncorrelated structure

To simulate trait-or tissue-specific variation unrelated to the predefined cluster structure, we added uncorrelated features to both *V*_*trait*_ and *V*_*tissue*_. We first ranked auxiliary traits and tissues by their total contribution to the simulated clusters, then randomly selected a subset of low-contributing (“non-important”) features. For each selected feature, we replaced its profile by resampling values from a top-contributing (“important”) feature of the same modality—maintaining realistic signal strength while disrupting alignment with the cluster structure. The number of such substituted traits and tissues defines the level of uncorrelated structure simulated (*s*).

#### Representative simulation example

We selected a representative simulation scenario with *K*_causal_ = 5 and *m* = 200 for visualization in **Fig. 2a**. To improve clarity, only 4 of the 5 clusters are shown, excluding the cluster with the weakest trait–tissue correlation based on *r*_proj_. To align colors between true and inferred clusters across methods, we computed correlations between the true and inferred trait and tissue profiles, assigning each inferred cluster the color of its best-matching true cluster.

### GWAS summary statistics

We applied partitioning methods to GWAS summary statistics for 165 focal diseases/traits (average of 290K samples and 77 fine-mapped loci), including 57 UK Biobank diseases/traits^41^ and 108 non-UK Biobank diseases/traits (**Supplementary Table 1**; see Data Availability).

#### Fine mapping

We conducted single causal variant fine-mapping on all tested GWAS traits as in ref ^20^.

#### Combining results across GWAS diseases/traits

To evaluate significant differences between J-PEP’s PEPA average across GWAS diseases/traits with respect to other partitioning strategies, we used two approaches. First, we applied a genomic-block jackknife to the unweighted average PEPA difference across traits (denoted *p* in the main text). Second, we performed a jackknife using trait-specific weights based on the inverse variance of each trait’s PEPA difference (denoted *p*_*w*_ in the main text).

### EpiMap data set

#### Track calling

We obtained EpiMap^36^ *p* value signal tracks from the Washington University Epigenome for downstream analysis. We restricted our selection to tracks that met two criteria: (1) the tracks were directly observed rather than imputed, and (2) the tracks corresponded to one of six epigenomic annotations — DNase hypersensitive sites (DNase HS), H3K4me1, H3K4me2, H3K4me3, H3K9ac, or H3K27ac. The selected tracks were initially downloaded in bigWig format and converted to bedGraph format using the UCSC command-line utility bigWigToBedGraph. Following this conversion, we performed peak calling using MACS2 to identify significant regions of epigenomic activity. Peaks were called using the macs2 bdgpeakcall command with a significance threshold of −log_10_(*p*) > 2, ensuring that only high-confidence peaks were included in the subsequent analyses.

#### SNP-to-tissue scoring using the EM algorithm

To compute SNP-to-tissue scores from epigenomic annotations, we applied an expectation-maximization (EM) algorithm to fine-mapped variants overlapping epigenomic peaks from the EpiMap project. SNPs were retained if their posterior inclusion probability (PIP) exceeded a threshold (default: 0.01). Functional annotations were filtered to exclude cancer samples and tracks with extreme background enrichment values (computed on chromosome 1 of the 1000 Genomes Project).

Let *Vϵ*{0,1}^*M*×*t*^ denote the binary overlap matrix of *M* SNPs and *t* epigenomic annotations. We initialize posterior weights *a*_*pt*_ across annotations for each SNP *p* by normalizing overlap rows:

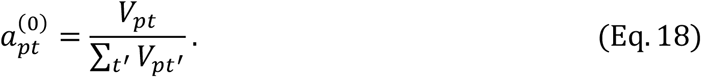

At each iteration, we re-estimate annotation weights *θ*_*t*_ as a weighted sum over SNPs:

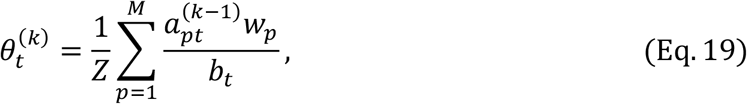

where *w*_*p*_ is the SNP-specific weight based on its PIP, *b*_*t*_ is the background expectation for annotation *t*, and *Z* is a normalization constant ensuring ∑_*t*_ *θ*_*t*_ = 1.

Posterior weights *a*_*pt*_ are then updated as:

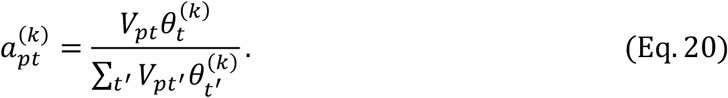

Iterations proceed until convergence is achieved (mean squared difference between 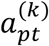 and 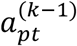 below 10^−8^, or a maximum of 200 iterations).

Annotation-level scores were aggregated into predefined tissue groups using EpiMap metadata^36^, yielding a final SNP-to-tissue matrix *V*_*tissue*_*ϵ*ℝ^*M*×*T*^, where *T* is the number of grouped tissues.

### Single-cell data set

We analyzed a single-cell atlas of chromatin accessibility comprising 615,998 cells across 111 fine-grained adult human cell types^42^. Cell-type–specific peak annotations were downloaded using the following command: *wget -r --level=1 -np -R index.html* http://catlas.org/catlas_downloads/humantissues/Peaks/.

We retained the original cell-type labels as provided by the authors. The SNP-to-cell-type association matrix *V*_*cell − type*_ was constructed using the same procedure applied to the bulk tissue annotations (see **EpiMap data set**) but substituting in the single-cell–derived peak tracks.

### Gene Ontology (GO) enrichment analysis

To investigate the biological processes associated with each cluster, we conducted Gene Ontology (GO) term enrichment analysis using the “Enrichr” R package. SNP-to-cluster scores were mapped to genes using pre-annotated SNP-to-gene mappings^44^, and gene-level scores were calculated by summing the scores of all SNPs assigned to each gene. To prioritize the most informative signals, genes were ranked by their cluster scores, and only the top genes cumulatively contributing to 80% of the total cluster weight were retained. GO terms were considered significant if they included at least four overlapping genes and had a Benjamini-Hochberg adjusted p-value below 0.05.

## Notes

### Competing Interest Statement

The authors have declared no competing interest.

### Author Declarations

The study exclusively used publicly available human data from open-access biobanks or research studies.

### Summary of Updates

Update version of the manuscript

