## Supplemental Material for "Mapping disease loci to biological processes via joint pleiotropic and epigenomic partitioning"

### Supplementary Note

##### Bounding the Reconstruction Error of $W$ in Pleiotropic Partitioning

We aim to show that the error in reconstructing the SNP-to-cluster membership matrix $W$ is lower when using pleiotropic data compared to epigenomic data under the **Pleiotropic partitioning** model. That is, we wish to prove:

$$L\big(W \mid V_{trait}, H_{trait}^{fit}\big) \le L\big(W \mid V_{tissue}, H_{tissue}^{fit}\big), \qquad \text{(Eq. 1)}$$

where $L$ indicates the reconstruction error $L(W \mid A, B) = \|A - WB\|_F$ (squared Frobenius norm), and:

1) $H_{trait}^{fit}$ is the trait profile matrix obtained from non-negative matrix factorization (bNMF) of $V_{trait}$, i.e.

$$\{W_{trait}, H_{trait}^{fit}\} = \underset{W\ge 0, H_{trait}\ge 0}{\operatorname{argmin}} \left\{\|V_{trait} - WH_{trait}\|^2 + \sum_k \lambda\left(W_k, H_{trait,k}\right)\right\}. \qquad \text{(Eq. 2)}$$

2) $H_{tissue}^{fit}$ is the least-squares estimate of tissue profiles from $V_{tissue}$ and the fixed $W_{trait}$, i.e.

$$\{H_{tissue}^{fit}\} = \underset{H_{tissue}\ge 0}{\operatorname{argmin}}\{\|V_{tissue} - W_{trait}H_{tissue}\|^2\} = ({W_{trait}}^T W_{trait})^{-1} {W_{trait}}^T V_{tissue}. \qquad \text{(Eq. 3)}$$

Let $W_{tissue} = \underset{W\ge 0}{\operatorname{argmin}}\left\{\left\|V_{tissue} - WH_{tissue}^{fit}\right\|^2\right\}$ be the best fit of W for reconstructing $V_{tissue}$. Then, using standard properties of least-squares projections and assuming without loss of generality that $V_{tissue}$ and $V_{trait}$ are unit-normalized,

$$\begin{aligned}
&L\big(W \mid V_{tissue}, H_{tissue}^{fit}\big) = \left\|V_{tissue} - W_{tissue}H_{tissue}^{fit}\right\|^2 \\
&= \left\|V_{tissue} - W_{tissue}({W_{trait}}^T W_{trait})^{-1}{W_{trait}}^T V_{tissue}\right\|^2 \\
&= \left\|(I_M - W_{tissue}({W_{trait}}^T W_{trait})^{-1}{W_{trait}}^T)V_{tissue}\right\|^2 \\
&= \left\|(I_M - W_{tissue}({W_{trait}}^T W_{trait})^{-1}{W_{trait}}^T)\right\|^2 \|V_{tissue}\|^2 \\
&= \left\|V_{trait} - W_{tissue}({W_{trait}}^T W_{trait})^{-1}{W_{trait}}^T V_{trait}\right\|^2 \approx \left\|V_{trait} - W_{tissue}H_{trait}^{fit}\right\|^2 \\
&\ge \left\|V_{trait} - W_{trait}H_{trait}^{fit}\right\|^2 \\
&= L\big(W \mid V_{trait}, H_{trait}^{fit}\big) \ \blacksquare
\end{aligned} \qquad \text{(Eq. 4)}$$

##### T2D results using the default set of 82 auxiliary traits

Applying J-PEP to T2D using the default set of 82 auxiliary traits (as opposed to the 109 auxiliary traits from ref. [25] described in the main text), we identified seven clusters. According to the PEPA metric, J-PEP (PEPA = 0.03) was not significantly different from either Epigenomic partitioning (PEPA = 0.02, $p$ = 0.09; 5 clusters) or Pleiotropic partitioning (PEPA = 0.03, $p$ = 0.45; 5 clusters). The resulting clusters were largely consistent with those reported in the main text and **Figure 5**, including epithelial, endocrine, liver, digestive, embryonic stem cell, and stromal-like clusters. Only one cluster differed markedly, exhibiting a distinct tissue profile with enrichment for brain-related signals (see *Data Availability* for numerical results).

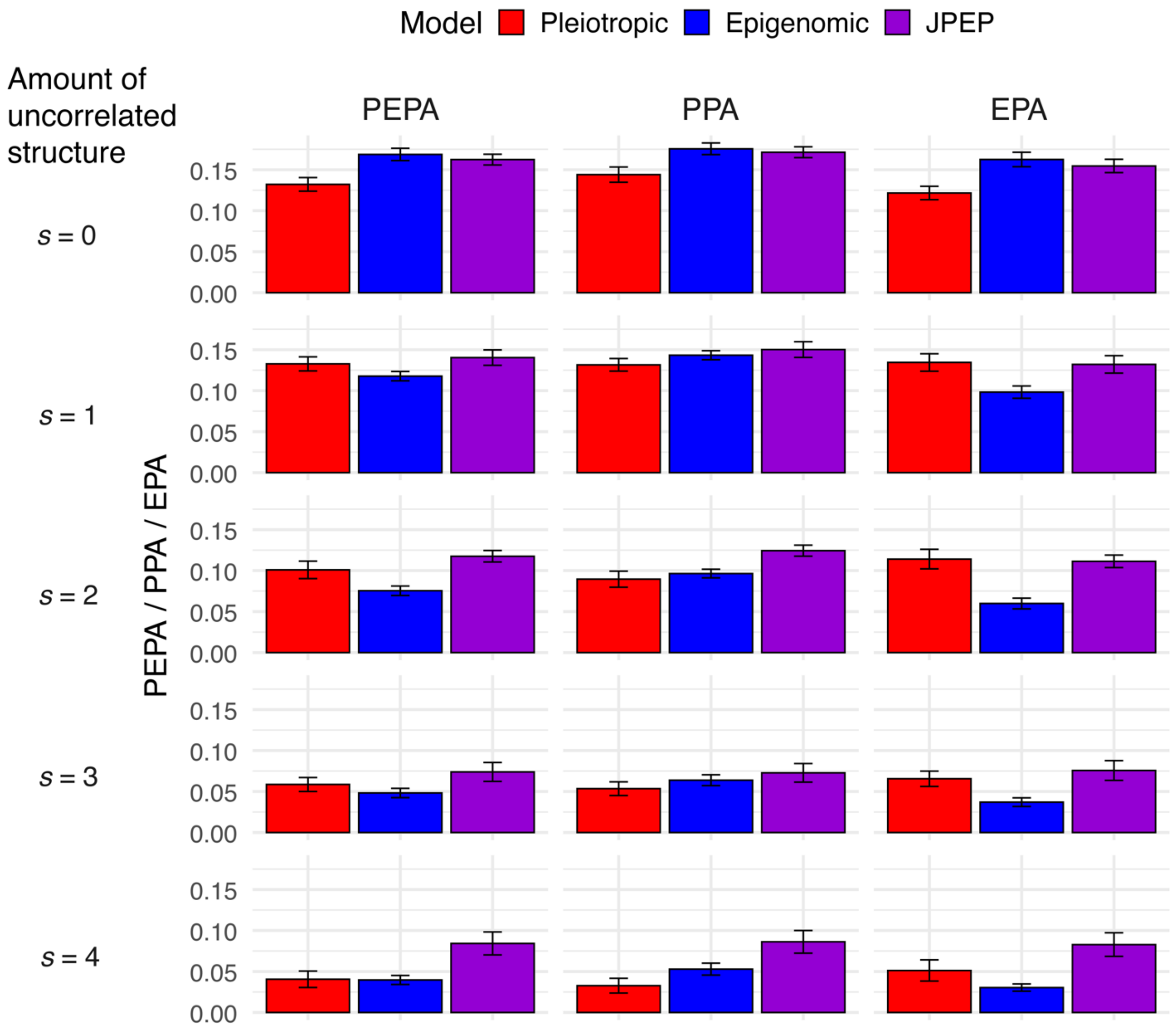


**Supplementary Fig. 1 | Prediction accuracy across models with varying levels of uncorrelated structure.**

Bar plots depict prediction accuracies PEPA, PPA and EPA for the Pleiotropic partitioning (red), the Epigenomic partitioning (blue) and J-PEP (purple) models across different levels of uncorrelated structure (rows, from none to 4 degrees). The y-axis represents mean prediction accuracy, with error bars denoting the standard error of the mean.

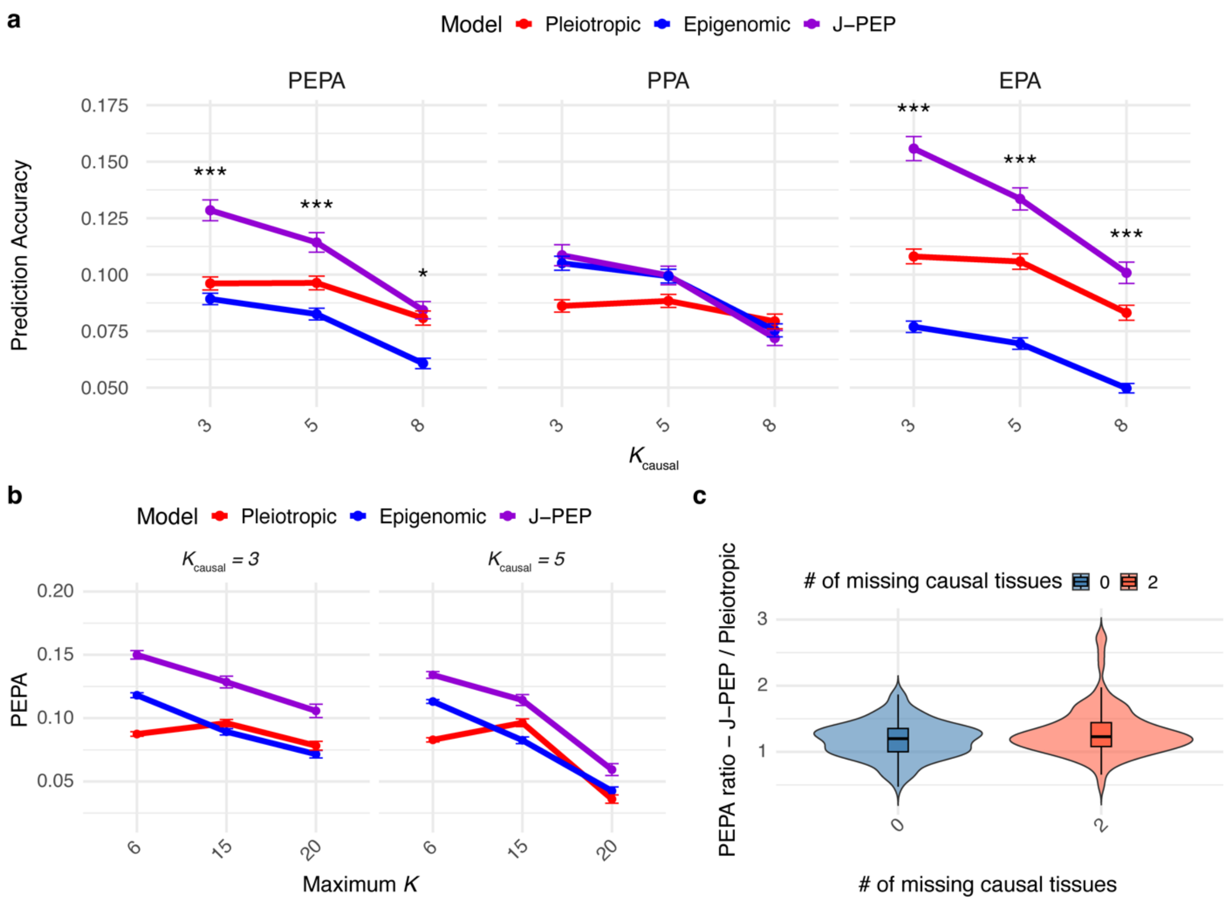


**Supplementary Fig. 2 | Simulations with varying parameters beyond uncorrelated structure.**

(a) Prediction accuracy for PEPA, PPA, and EPA metrics across 100 simulation replicates with varying numbers of causal clusters ($K_{causal}$ = 3, 5, 8) and $m$ = 400 causal SNPs for the focal disease/trait. Error bars denote standard errors; significance levels: *: $p < 0.05$, **: $p < 0.01$, ***: $p < 0.001$. (b) PEPA scores across 100 replicates with varying maximum allowed clusters ($K$ = 6, 15, 20), two $K_{causal}$ values (3 and 5), and $m$ = 400 causal SNPs for the focal disease/trait. (c) Distributions of PEPA ratios (J-PEP vs. Pleiotropic partitioning) in scenarios with 0 (blue) or 2 (red) missing causal tissues, for $K_{causal}$ = 5 and $m$ = 400 causal SNPs for the focal disease/trait.

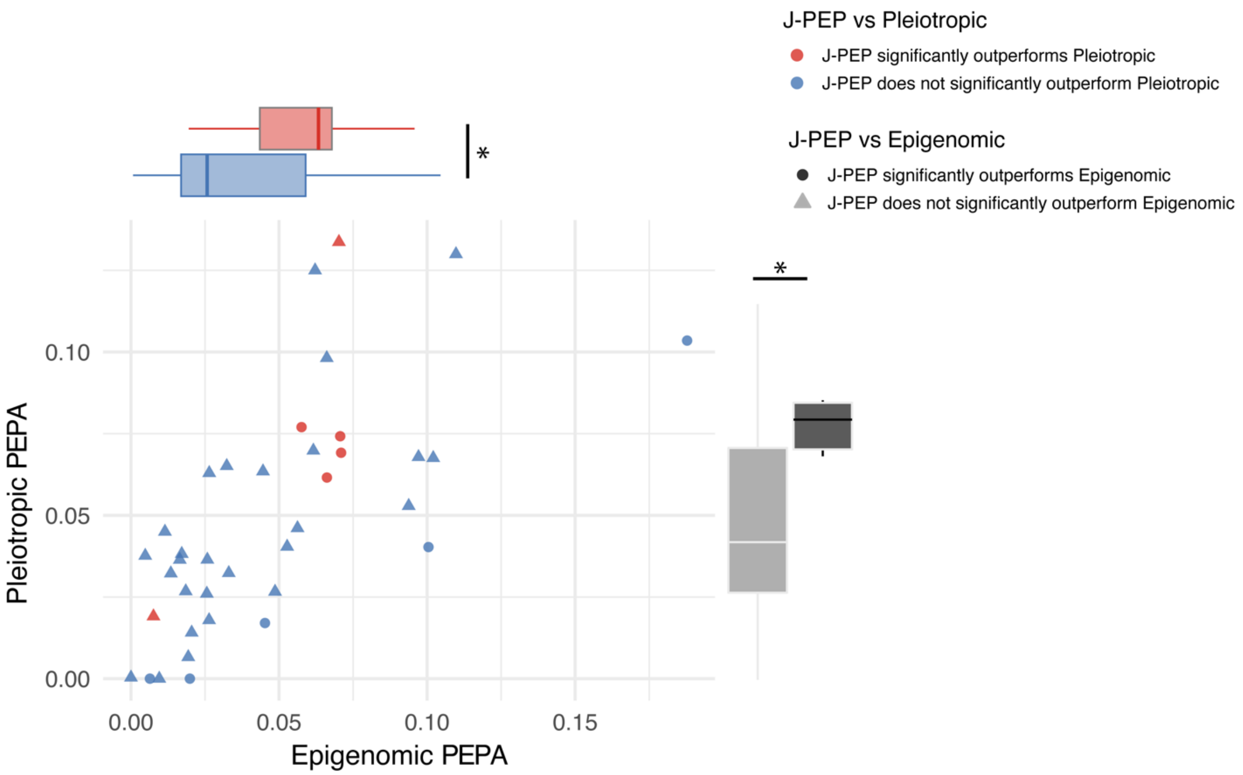


**Supplementary Fig. 3 | J-PEP Integrates Complementary Information from Pleiotropic and Epigenomic Partitioning.**

Scatter plot shows PEPA values for the 38 focal traits under Epigenomic partitioning (x-axis) and Pleiotropic partitioning (y-axis). Each dot represents a focal trait, with colors denoting J-PEP's performance relative to Pleiotropic partitioning and shapes indicating J-PEP's performance relative to Epigenomic partitioning. Marginal boxplots illustrate the distribution of PEPA values along each axis, grouped by J-PEP's comparative performance against Pleiotropic (x-axis) and Epigenomic (y-axis).

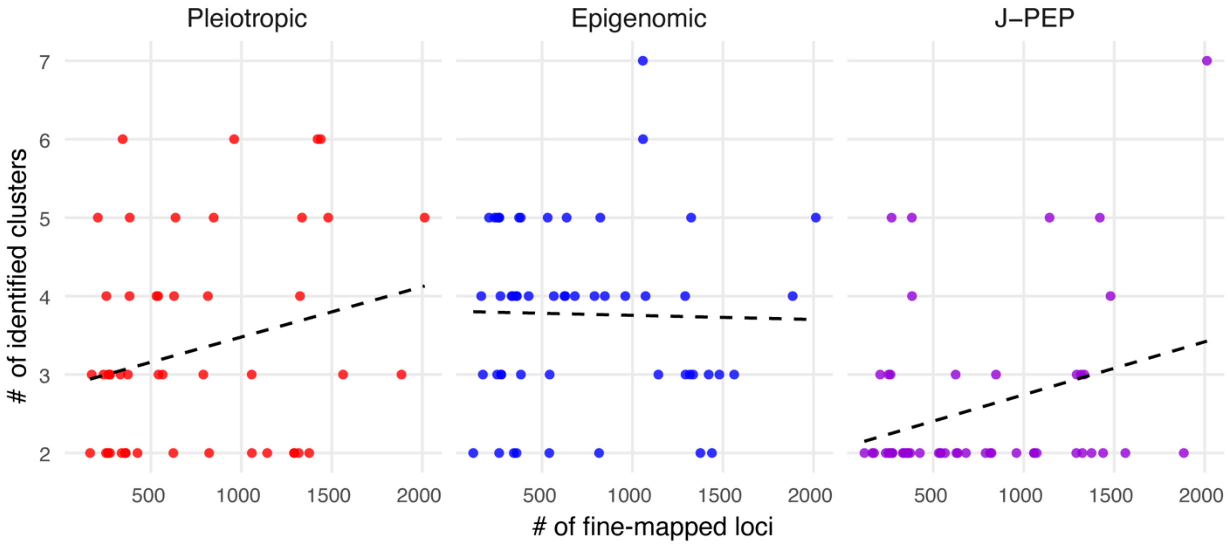


**Supplementary Fig. 4 | The number of identified clusters correlates with GWAS power.** Scatterplots show the relationship between the number of clusters identified per focal trait (y-axis) and the number of fine-mapped loci (x-axis) across focal traits for each clustering method: Pleiotropic partitioning (red), Epigenomic partitioning (blue), and J-PEP (purple). Each point represents one focal trait.

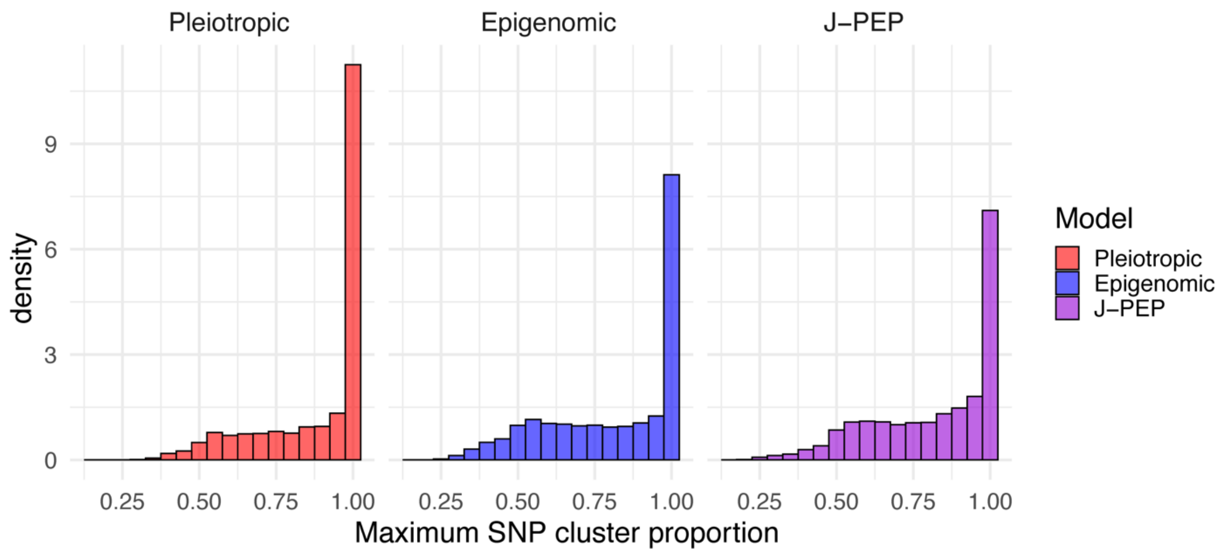


**Supplementary Fig. 5 | Distribution of SNP cluster purity across models.**
Histograms show the distribution of maximum SNP cluster proportions for the Pleiotropic partitioning (red), Epigenomic partitioning (blue), and J-PEP (purple) models. The x-axis represents the highest proportion of assignment for each SNP to a single cluster, while the y-axis denotes the density of SNPs. Higher values on the x-axis indicate stronger SNP-to-cluster differentiation. In addition, we computed a concordance rate at each locus-focal trait pair, representing the average maximum normalized sum of cluster membership across SNPs (see main text).

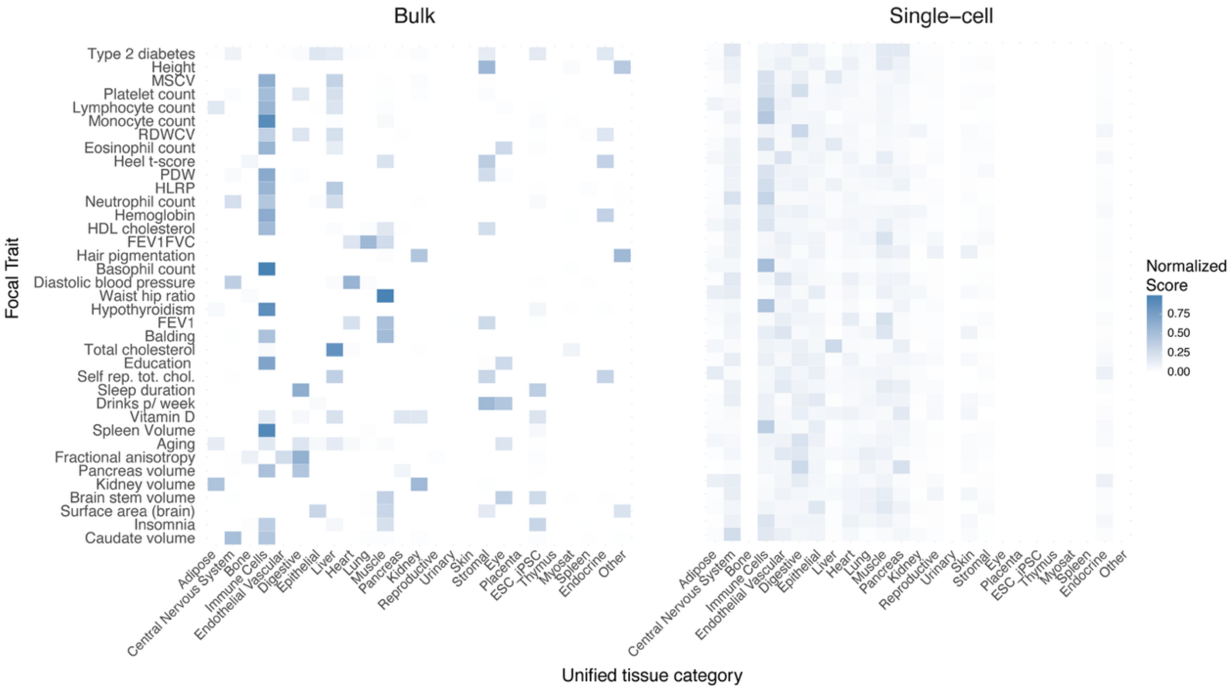


**Supplementary Fig. 6 | Tissue and cell-type–trait association heatmap across epigenomic modalities.**

The heatmap shows the aggregated tissue-to-cluster scores for each of the 38 focal traits (rows) across unified tissue categories (columns; see **Supplementary Table 7**), separately for bulk (left) and single-cell (right) epigenomic annotations. Scores represent the normalized sum of tissue contributions across a given focal trait's clusters. abb. MSCV: Mean sphered corpuscular volume; RDWCV: Red cell distribution width - coefficient of variation; PDW: Platelet distribution width; HLRP: High light reticulocyte proportion; FEV1: Forced expiratory volume in 1 second; FEV1FVC: Forced expiratory volume in 1 second/forced vital capacity.

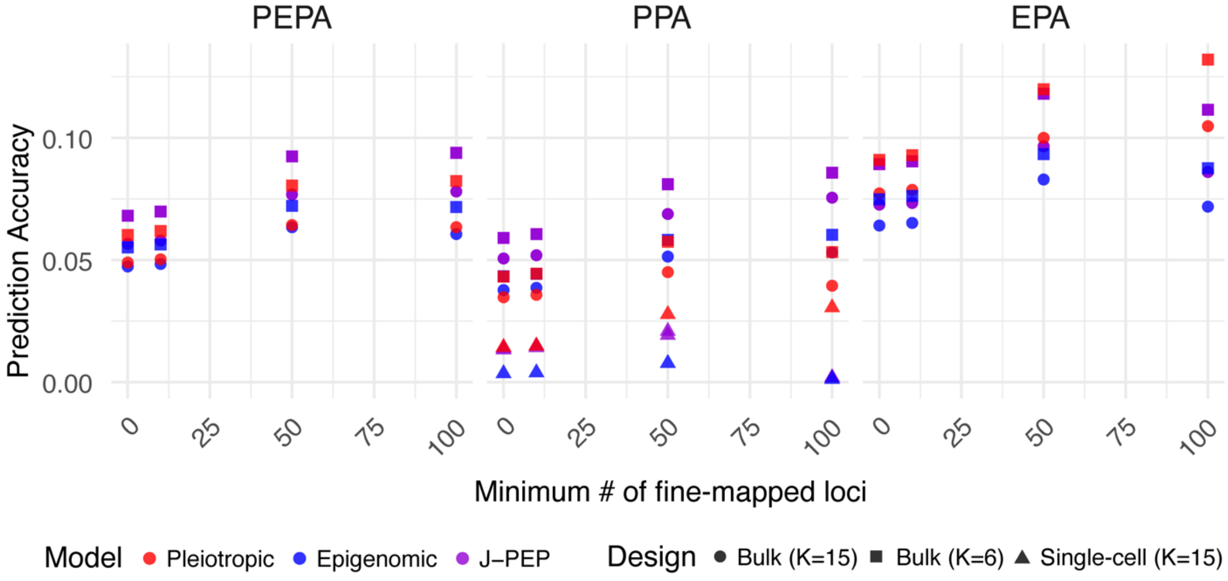


**Supplementary Fig. 7 | J-PEP maintains superior performance across cluster settings, while single-cell–derived clustering underperforms.**
Prediction accuracy metrics PEPA (left), PPA (center), and EPA (right) as a function of the minimum number of fine-mapped loci. Colors indicate models: Pleiotropic partitioning (red), Epigenomic partitioning (blue) and J-PEP (purple). Shapes indicate the clustering design: K = 15 using bulk data (circle), K = 6 using bulk data (square), and K = 15 using single-cell data (triangle). Single-cell results are shown only for PPA due to non-comparable dimensionality across metrics resulting from differing cell-type and tissue annotations across epigenomic modalities.

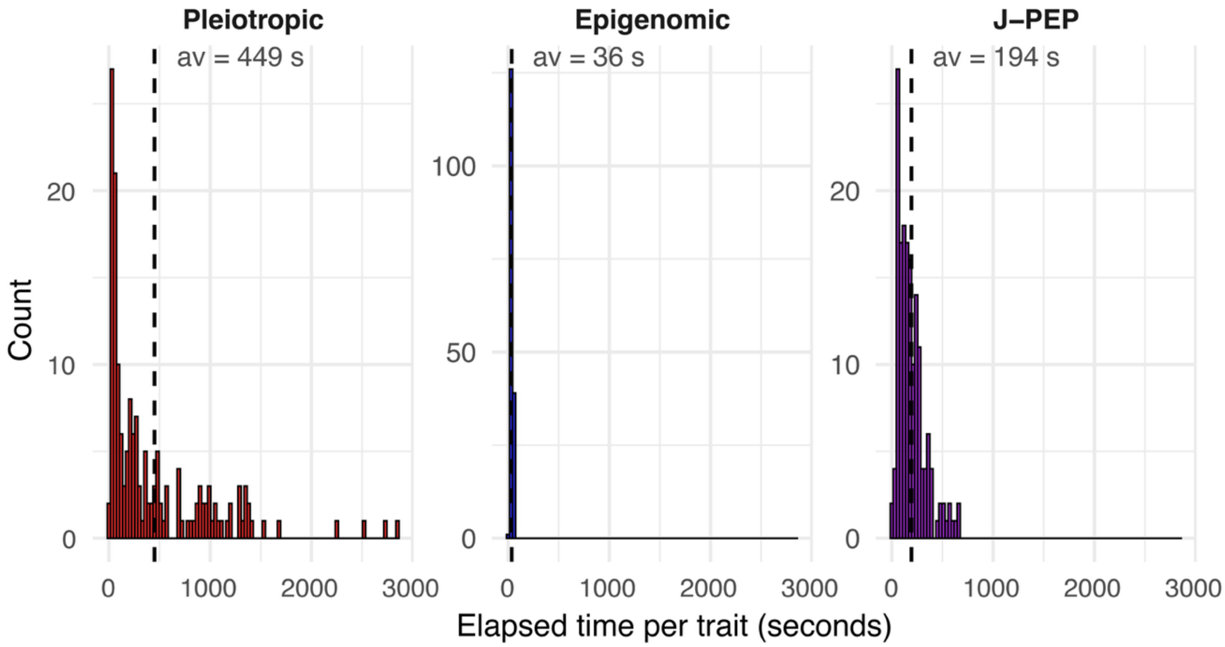


**Supplementary Fig. 8 | Runtime comparison across partitioning methods.**
Distribution of runtimes (in seconds) for Pleiotropic partitioning, Epigenomic partitioning, and J-PEP across all 165 traits. Vertical dashed lines indicate the mean runtime per method. Reported times reflect the total duration of ten independent runs per trait-method combination, as required by the default optimization procedure. We note that J-PEP had lower running times than Pleiotropic partitioning, likely due to the smaller size of epigenomic data vs. pleiotropic data and the focus of J-PEP in shared structure.

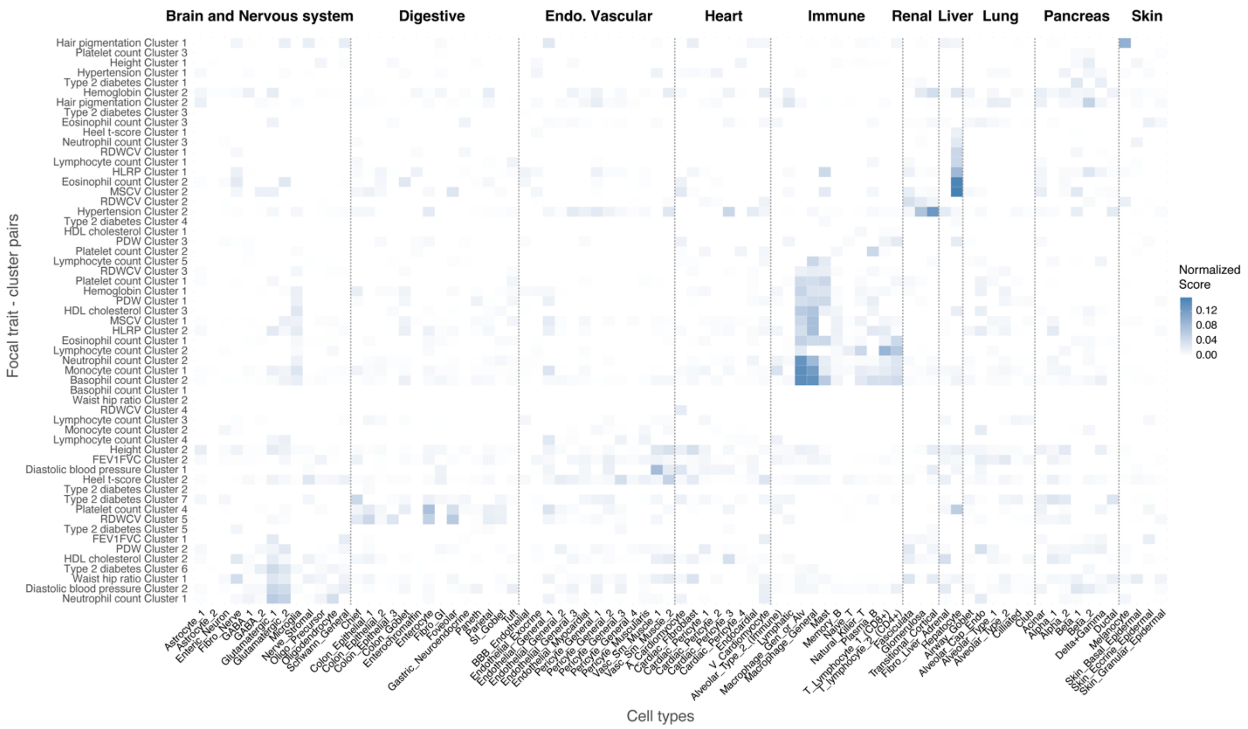


**Supplementary Fig. 9 | Cell-type association heatmap across focal trait-cluster pairs.** This heatmap displays normalized cell-type scores for all trait-cluster pairs associated with the 20 focal traits shown in **Fig. 3a**. Rows represent trait-cluster combinations, and columns correspond to 24 single-cell-derived cell types grouped by unified tissue categories (top, see **Supplementary Table 7**), including immune, heart, renal, and liver. Scores indicate the proportion of each cluster linked to a given cell type, normalized across all cell types for that cluster. This figure provides an extension of **Fig. 4**. abb. MSCV: Mean sphered corpuscular volume; RDWCV: Red cell distribution width - coefficient of variation; PDW: Platelet distribution width; HLRP: High light reticulocyte proportion; FEV1FVC: Forced expiratory volume in 1 second/forced vital capacity.

#### Supplementary Tables (separate excel files)

**Supplementary Table 1. Summary of GWAS diseases/traits.**
The column `Approx. genet. indep.` denotes whether a trait was part of the set of 55 approximately genetically independent traits selected for benchmarking analyses. The column `Criteria met` identifies the subset of 38 traits for which at least two robust clusters (see **Methods**) were identified consistently across all three partitioning methods: Pleiotropic partitioning, Epigenomic partitioning, and J-PEP. Note that `All_Metal_LDSC-CORR_Neff` denotes the trait T2D when analyzed with the default set of 82 auxiliary traits from this work, whereas `All_Metal_LDSC-CORR_Neff_comparison` denotes the trait T2D when analyzed with the set of auxiliary traits from ref. [25]. In both cases, we use the same summary statistics for T2D.

**Supplementary Table 2. Simulation benchmarking: Frobenius norm errors, and PEPA values.**

**Supplementary Table 3. PEPA metric values across 165 diseases/traits for J-PEP, Pleiotropic, and Epigenomic partitioning.**
Missing values indicate cases where prediction was not performed because fewer than two clusters were identified in at least 5 out of the 22 chromosome-specific runs. Number of clusters is indicated for each trait-model pair (K_JPEP, K_EPI, K_PLEIO). The column `Criteria met` identifies the subset of 38 traits for which at least two robust clusters (see **Methods**) were identified consistently across all three partitioning methods: Pleiotropic partitioning, Epigenomic partitioning, and J-PEP. Note that `All_Metal_LDSC-CORR_Neff` denotes the trait T2D when analyzed with the default set of 82 auxiliary traits from this work, whereas `All_Metal_LDSC-CORR_Neff_comparison` denotes the trait T2D when analyzed with the set of auxiliary traits from ref. [25]. In both cases, we use the same summary statistics for T2D.

**Supplementary Table 4. Projection correlation scores across 38 diseases/traits.**
Average across clusters for each focal trait.

**Supplementary Table 5. Epigenomic tracks used in bulk-level analyses.**
Number of tissue-specific epigenomic tracks and tissue-to-cluster scores for focal trait-model pairs are shown.

**Supplementary Table 6. Single-cell–derived cluster profiles across 38 diseases/traits.**
Single-cell–derived cluster profiles are shown sequentially and were obtained using the SNP-to-cluster membership matrix $W$ derived from the respective partitioning method using bulk-level data. Note that `All_Metal_LDSC-CORR_Neff` denotes the trait T2D when analyzed with the default set of 82 auxiliary traits from this work, whereas `All_Metal_LDSC-CORR_Neff_comparison` denotes the trait T2D when analyzed with the set of auxiliary traits from ref. [25]. In both cases, we use the same summary statistics for T2D.

**Supplementary Table 7. Cell type–to–tissue mapping for single-cell chromatin accessibility data.**
$H_{tissue}$ matrices obtained from single-cell data are sequentially shown for different focal traits.

**Supplementary Table 8. SNP-to-trait matrices for T2D, HTN and NC analyses.**
$V_{trait}$ matrices are shown one after the other for T2D (as presented in the main text), HTN and NC.

**Supplementary Table 9. GO enrichment results for clusters in T2D, HTN and NC.**
Significant GO terms for T2D (as presented in the main text), HTN and NC clusters. GO terms were considered significant if they included at least four overlapping genes and had a Benjamini-Hochberg adjusted $p$-value below 0.05.
